# Acceptability and feasibility of measles vaccination, small-quantity lipid-based nutrient supplementation, and their co-delivery among children aged 6–23 months in northern Nigeria: a qualitative study

**DOI:** 10.64898/2026.11.06.26367420

**Authors:** Saidu Idris Ahmad, Juliet Nabunje, Samson Kumzhira Adamu, Aisha Muhammad Sheriff, Abdulganiyu Gimba Gwayo, Aisha Goje, Irine Atanga, Ariong’o M Akinyi, Emeka Chikezie Ugonna, Soma Bahonon, Umar Chiroma, Hadiza Maina Adam, Renaud Becquet, Nuru Suleiman Muhammad, Baba Waru Goni, Cécile Cazes, Kevin P.Q. Phelan, Mélanie Plazy

**Author notes:** **Corresponding author** Melanie Plazy.

## Abstract

**Background:** Undernutrition and infectious diseases form a vicious cycle that endangers child survival in low-resource settings, including in Northern Nigeria. The NutriVax-Measles trial was designed to evaluate a strategy combining preventive small-quantity lipid-based nutrient supplements (SQ-LNS) distributions with community mobilization and routine immunization services for children aged 6-23 months to improve measles vaccination uptake. To inform future programs co-delivering nutrition supplementation and immunization services at scale, we aimed to assess the feasibility and acceptability of measles vaccination, SQ-LNS distribution, and the NutriVax strategy.

**Methods:** A qualitative study was conducted at the end of the trial using in-depth interviews (n=53) and focus group discussions (n=8) with key stakeholders: caregivers, health care workers, community health workers, and community representatives. Thematic analysis was guided by multiple conceptual frameworks.

**Results:** The majority of the participants expressed satisfaction with measles vaccines in both arms, and with SQ-LNS in the NutriVax arm, as they perceived the benefits for their children’s health. In both arms, community awareness sessions and previous experience of outbreaks were key to motivating caregivers to bring their children to healthcare centres. Although distance to health facilities was identified as one of the main barriers to accessing care, SQ-LNS was perceived as an important source of motivation to bring their child to the clinic where they subsequently received vaccination. All stakeholder groups emphasized the importance of community leaders’ support and targeted male sensitization to address the prevailing sociocultural norms. Additionally, improved service organization to reduce waiting time was considered crucial to enhance caregivers’ acceptability of the intervention.

**Conclusion:** Both measles vaccination and SQ-LNS provision were acceptable and feasible, supporting the NutriVax strategy within primary healthcare settings in fragile health systems. Successful implementation depends on sustained community sensitization, continued engagement of fathers and community leaders, and service delivery decentralized as close as possible to beneficiaries.

**KEY MESSAGES:** *What is already known?:*

- Co-delivering small-quantity lipid-based nutrient supplements (SQ-LNS) with routine immunization is an effective strategy to improve vaccination coverage in Northern Nigeria. Yet, evidence on how vaccination and SQ-LNS are perceived, implemented, and experienced by stakeholders at health-care and community levels remains scarce.

*What are the new findings?:*

- SQ-LNS was an important source of motivation for caregivers to bring their children to health facilities, creating opportunities for vaccination.
- Persistent barriers to vaccination included socio-cultural misconceptions, gender norms, long waiting time and distance to health facilities.

*What do the new findings imply?:*

- Sustained community engagement, including targeted male sensitization, is important to improve measles vaccination acceptability.
- Successful implementation of co-delivery programs also requires well-organized service delivery, with services decentralized to health facilities as close as possible to beneficiaries, particularly to reduce travel distances and waiting times.

## Introduction

Undernutrition and infectious diseases are interlinked in a vicious cycle that continues to threaten child survival (1). Undernutrition weakens immune function and increases children’s susceptibility to infectious diseases, while vaccine-preventable infections, such as measles, further worsen nutritional status, increasing the risk of mortality and poor developmental outcomes (2–4). Sub-Saharan Africa bears a disproportionate share of this dual burden (5), with Nigeria among the countries with both the highest prevalence of undernutrition and the lowest vaccination coverage worldwide, positioning it at the epicentre of the undernutrition–infectious disease cycle. In 2023, 34.2% of children aged 6-59 months were stunted and 6.5% were wasted nationwide (6), with a high burden of undernutrition-related child mortality (5). The country also experiences recurrent measles outbreaks, particularly in northern states (7), and measles contributes to an under-five mortality rate of 105 per 1,000 live births, or roughly 768,000 childhood deaths annually, far above the Sustainable Development Goals target of 25 per 1,000 by 2030 (8). Despite immunization being one of the most effective public health interventions for preventing child mortality (9,10), more than 2.2 million children were unvaccinated countrywide in 2021 (11). Multiple barriers limit vaccination uptake in low-and middle-income countries, including weaknesses in health systems, vaccine availability, caregivers’ education, and socio-cultural misconceptions (12). Evidence on strategies to improve childhood vaccination coverage in low- and middle-income countries remains limited, with little or modest effects of health education, SMS reminders, or financial incentive approaches (13). Meanwhile, small-quantity lipid-based nutrient supplements (SQ-LNS) have emerged as an effective intervention to improve child nutrition, reducing severe wasting, stunting, and all-cause mortality in young children (14,15), leading the World Health Organization to recommend their use in certain food-insecure settings, including priority countries such as Nigeria (16). These complementary benefits suggest that co-delivering preventive nutrition supplements with routine immunization could simultaneously address undernutrition and low vaccination coverage, yet evidence on such integrated strategies remains scarce.

The NutriVax-Measles trial showed that a strategy combining SQ-LNS supplementation and routine immunization services for children aged 6-23 months significantly increased measles vaccination uptake in Northern Nigeria (17). Beyond demonstrating the effectiveness of this intervention, understanding how measles vaccination and SQ-LNS are perceived, implemented, and experienced by different stakeholders is important to inform future programs co-delivering nutrition supplementation and immunization services at scale (18).

The objective of this qualitative study was to assess the feasibility and acceptability of measles vaccination, SQ-LNS distribution, and the NutriVax strategy in Northern Nigeria and to identify factors that could inform future scale-up.

## Methods

### Study design

This study employed an exploratory qualitative research design nested in the NutriVax-Measles trial, following SRQR guidelines (Supplementary file 1). Semi-structured in-depth interviews (IDIs) and focus group discussions (FGDs) were conducted from 16-May to 30-July 2025 among various stakeholders in both health system (healthcare workers, community health workers) and community (parents/caregivers and community representatives).

### Study setting: the NutriVax-Measles trial

The NutriVax-Measles study was a pediatric pragmatic superiority parallel cluster-randomized controlled trial conducted in Yobe State, Northeastern Nigeria. The main objective was to evaluate an innovative intervention designed to improve measles vaccination coverage among children aged 6-23 months by distributing SQ-LNS alongside routine vaccinations at primary health care centers (Figure 1). The intervention was implemented from July 2024 to July 2025 in Karasuwa and Nguru Local Government Areas (LGAs): each LGA comprised ten wards (clusters), five assigned for the intervention arm (NutriVax arm) and five for the control arm. A comprehensive study protocol detailing all study objectives, eligibility criteria, and methodological approach has been published previously (19). All clusters followed the National Programme on Immunization (NPI) schedule, providing routine vaccination activities at Primary Health Care Centres (PHCCs) or Health Posts (HPs), with vaccines administered by Ministry of Health (MoH) staff according to MoH plans. Community engagement and mobilization were provided by Community Nutrition Mobilizers (CNMs) at health facilities and within communities, to sensitize caregivers and promote vaccination, infant and young child feeding and dietary diversity following standardized procedures. In the NutriVax clusters, caregivers of children aged 6-23 months received additional messages on SQ-LNS eligibility, and that they could receive monthly rations of one sachet of SQ-LNS per day for up to one year or until reaching 24 months of age, at the PHCC on specific session days.

**Figure 1.**
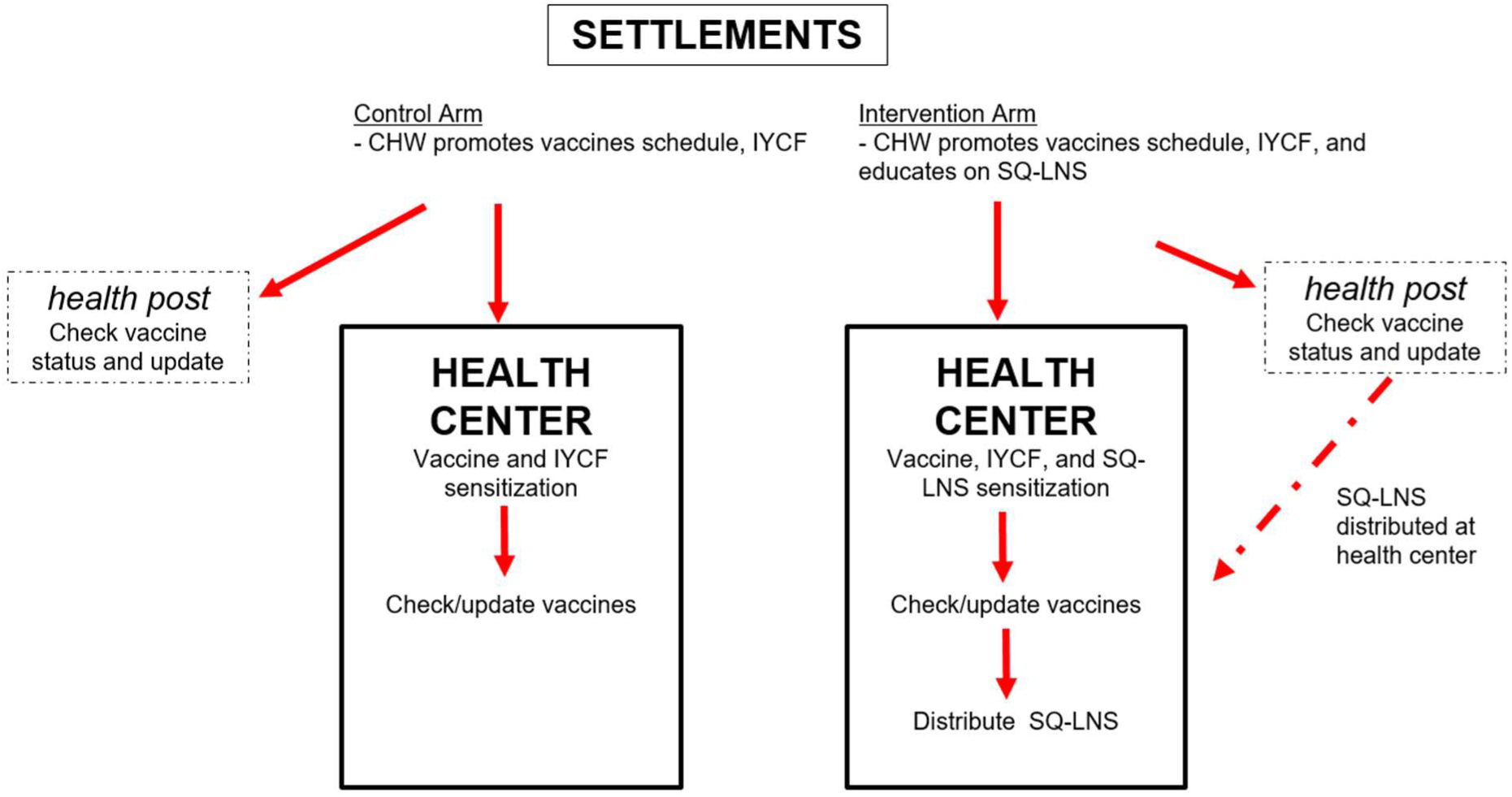
Description of the NutriVax-Measles intervention compared to the standard care CHW: community health workers; IYCF Infant and Young Children Feeding practices; SQ-LNS Small quantity lipid-based nutritional supplements

Several adaptations to intervention implementation were introduced during the trial, including: improving the SQ-LNS-vaccination circuit to reduce waiting times; improve the comfort of waiting areas with the construction of shaded areas; forming men’s groups and engaging community and religious leaders to increase awareness of the benefits of vaccination in both trial arms and of SQ-LNS in the intervention arm; working with MoH authorities to improve the availability of paracetamol for managing post-vaccination side effects; and providing up to three-months of SQ-LNS rations to reduce the frequency of visits for caregivers living far from PHCCs.

### Study population and sampling strategy

This qualitative study was conducted in 12 of the 20 wards included in the NutriVax-Measles trial, with six wards purposively selected from each LGA (three in the NutriVax arm and three in the control arm). Wards were selected to capture variation in health centers access (Supplementary file 2). Four stakeholder groups participated in the study: (1) parents or guardians of children aged 6-23 months, (2) health care workers (HCWs), (3) community health workers (CHWs), and (4) community representatives. The qualitative study included IDIs with participants from each stakeholder group across all selected wards in both trial arms, as well as two FGDs per stakeholder group, with one conducted in each LGA, including participants from intervention wards only. Participants were purposively selected according to eligibility criteria and predefined characteristics for each stakeholder group to ensure diversity in participant profiles and capture a broad range of perspectives and experiences within the NutriVax-Measles trial (Table 1).

**Table 1.**
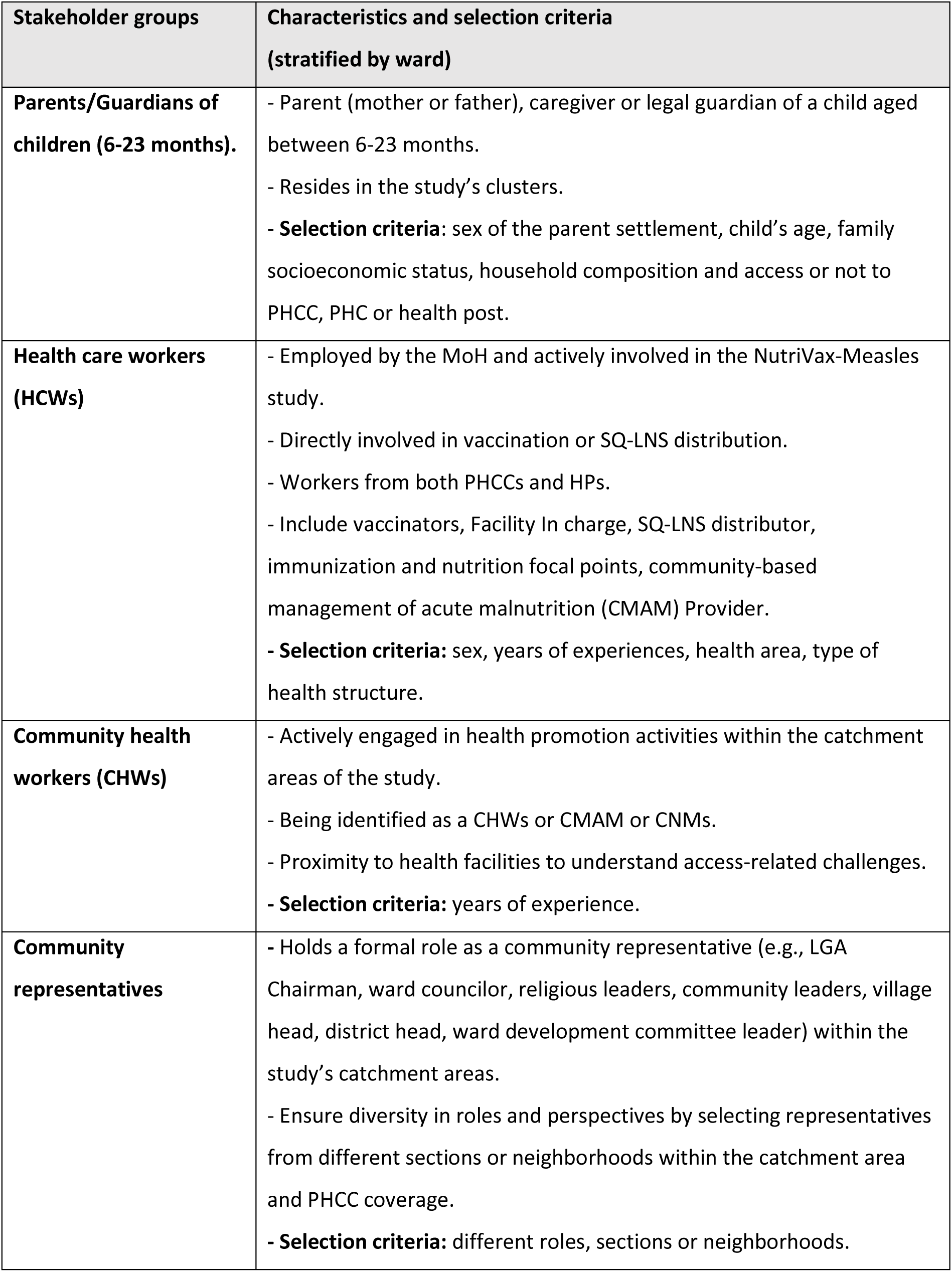
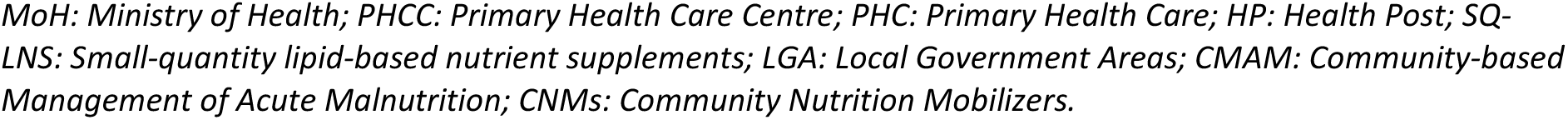
Stakeholder groups and selection criteria.

| Stakeholder groups | Characteristics and selection criteria<br>(stratified by ward) |
|---|---|
| **Parents/Guardians of children (6-23 months).** | - Parent (mother or father), caregiver or legal guardian of a child aged between 6-23 months.<br>- Resides in the study’s clusters.<br>- **Selection criteria**: sex of the parent settlement, child’s age, family socioeconomic status, household composition and access or not to PHCC, PHC or health post. |
| **Health care workers (HCWs)** | - Employed by the MoH and actively involved in the NutriVax-Measles study.<br>- Directly involved in vaccination or SQ-LNS distribution.<br>- Workers from both PHCCs and HPs.<br>- Include vaccinators, Facility In charge, SQ-LNS distributor, immunization and nutrition focal points, community-based management of acute malnutrition (CMAM) Provider.<br>**- Selection criteria:** sex, years of experiences, health area, type of health structure. |
| **Community health workers (CHWs)** | - Actively engaged in health promotion activities within the catchment areas of the study.<br>- Being identified as a CHWs or CMAM or CNMs.<br>- Proximity to health facilities to understand access-related challenges.<br>**- Selection criteria:** years of experience. |
| **Community representatives** | **-** Holds a formal role as a community representative (e.g., LGA Chairman, ward councilor, religious leaders, community leaders, village head, district head, ward development committee leader) within the study’s catchment areas.<br>- Ensure diversity in roles and perspectives by selecting representatives from different sections or neighborhoods within the catchment area and PHCC coverage.<br>**- Selection criteria:** different roles, sections or neighborhoods. |

*MoH: Ministry of Health; PHCC: Primary Health Care Centre; PHC: Primary Health Care; HP: Health Post; SQ-LNS: Small-quantity lipid-based nutrient supplements; LGA: Local Government Areas; CMAM: Community-based Management of Acute Malnutrition; CNMs: Community Nutrition Mobilizers.*

### Data collection and conceptual frameworks

Semi-structured interview guides (Supplementary file 3) were developed for each stakeholder group based on five conceptual frameworks: (1) WHO’s Behavioral and Social Drivers (BeSD) framework (20), to explore vaccination uptake, (2) the Andersen’s Model (21) and (3) the Levesque’s framework (22) to explore access to care, (4) the Peh’s adherence framework (23) to investigate the use of the SQ-LNS, and (5) the Consolidated Framework for Implementation Research (CFIR) (24) to identify the main barriers and facilitators of the acceptance and feasibility of implementation at several levels (individual, social, health-system, structural). In addition, a gender lens was considered during development of the guides, to explore how gender norms, household roles, and decision-making dynamics influenced care-seeking behavior, vaccination uptake, and participation in SQ-LNS distribution.

The interview guides were first drafted in English, translated into Hausa by a professional translator, and pre-tested to ensure contextual and linguistic clarity. Thematic saturation was assessed during weekly analytical debriefings, during which emerging themes were reviewed across completed interviews and discussions. Data collection continued until these weekly reviews indicated that no new themes or insights were emerging from additional interviews or discussions.

All qualitative data were collected by a team of four trained qualitative research assistants (SAK, AMS, AGG, AG), all from Northern Nigeria, under the supervision of the social science research manager, also Nigerian (AS) and a French senior researcher (MP).

### Data analysis

A rapid qualitative thematic analysis approach (25,26) was used to explore the acceptability and feasibility of measles vaccination, SQ-LNS distribution, and the NutriVax intervention. This approach comprised several steps to enable timely synthesis of insights: (1) each interview was conducted by a team of two research assistants: one serving as the interviewer and the other one as the note-taker. A thematic summary grid template (27), specific to each stakeholder group was used to summarize key enablers, barriers, and adaptations across themes and subthemes simultaneously during participant interview (Supplementary file 4); (2) following each day of fieldwork, debriefing meetings were conducted between the social science research manager (SIA) and the qualitative research assistants, during which the team completed a global summary matrix organized by stakeholder group; (3) weekly debriefing meetings were conducted between the social science research manager (SIA), the NutriVax research manager (JN), and a senior researcher (MP) to review completed matrices, discuss patterns across stakeholder groups, and refine interpretations; (4) the conceptual frameworks (Supplementary file 5) guided the systematic and contextually grounded interpretation of findings, which were subsequently organized at the individual, social, health-system and structural levels; (5) audio recordings were transcribed and simultaneously translated from Hausa to English, allowing the transcripts to complement the rapid qualitative analysis with relevant illustrative quotations. Findings were discussed and validated with the research and operational teams during regular project meetings. Figure 2 presents an overview of the data analysis process.

**Figure 2.**
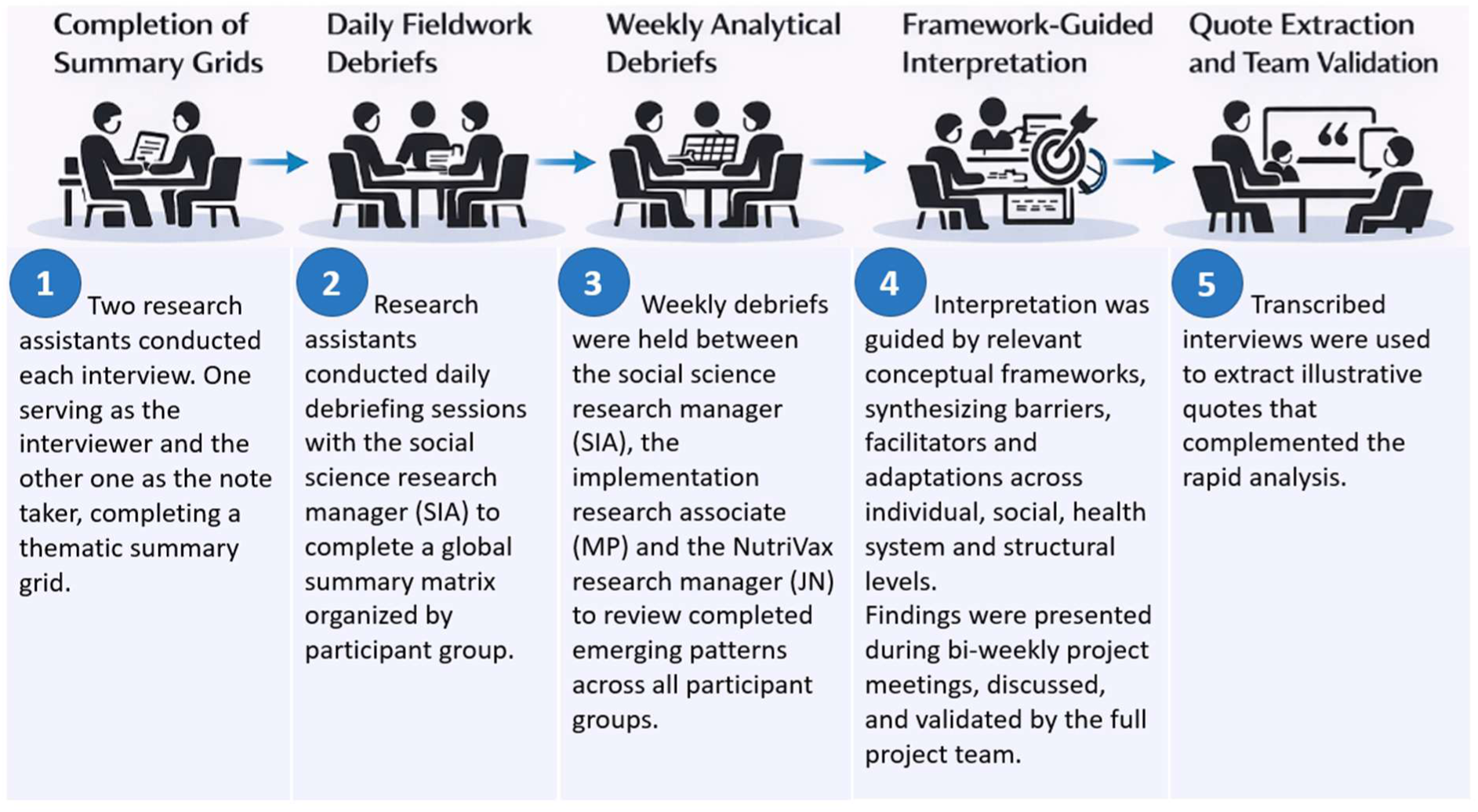
Rapid qualitative data collection and analysis process

### Patient and public involvement

As part of the NutriVax-measles study, community engagement and adherence were fostered from the preparation phase through meetings with community representatives and local Ministry of Health (MoH) nutrition and immunisation stakeholders. Throughout the trial, joint supervisory visits involving local representatives and Nigerian co-investigators monitored protocol adherence and ensured that implementation remained responsive to the local context. Engagement was also supported by a national steering committee bringing together MoH representatives at federal, state and local levels, nutrition and immunization partners, and community leaders. Several stakeholders, including community members, contributed in this research by participating in in-depth interviews and FGDs. Preliminary results of this work were presented at local and national levels, with the participation of community and MoH representatives.

### Ethics

Ethical approval was obtained from the Yobe State Ministry of Health and Human Services Health Research Ethics Committee (YOHREC) on February 14^th^, 2024 (MOH/GEN/747_February 2024) and from the Ethics Evaluation Committee of the French National Institute for Health and in France from the National Institute for Health and Medical Research (Inserm) Ethics Evaluation Committee (24-1091-20240402/IRB0000388). Written informed consent was obtained from all participants before the interview. Participants were informed about the purpose of the study and their right to withdraw at any time without any consequences. Recordings were stored on a secure drive accessible only to the qualitative research team.

## Results

### Description of the study participants

At the end of the trial, a total of 104 participants were involved in the study: 53 individuals participated in IDIs and 51 participated in 8 FGDs (Table 2). All community representatives were male, while females predominated in the other stakeholder groups.

**Table 2.** Characteristics of study participants - Final qualitative evaluation of the NutriVax trial.

|  | **Parents/legal guardian** | **Community representative** | **Community health workers** | **Healthcare workers** |
|---|---|---|---|---|
| **IDI (both arms)** | **n=16** | **n=12** | **n=13** | **n=12** |
| **Sex** |  |  |  |  |
| Female | 14 | 0 | 11 | 9 |
| Male | 2 | 12 | 2 | 3 |
| **Age** |  |  |  |  |
| Median (IQR) | 39 (37-49) | 49 (47-49) | 29 (21-29) | 39 (29 -42) |
| **LGA (arm)** |  |  |  |  |
| Nguru (NutriVax) | 6 | 2 | 2 | 2 |
| Nguru (Control) | 5 | 4 | 4 | 5 |
| Karasuwa (NutriVax) | 3 | 3 | 5 | 3 |
| Karasuwa (Control) | 2 | 3 | 2 | 2 |
| **FGD (intervention arm)** | **n=13** | **n=12** | **n=12** | **n=14** |
| **Sex** |  |  |  |  |
| Female | 11 | 0 | 10 | 10 |
| Male | 2 | 12 | 2 | 4 |
| **Age** |  |  |  |  |
| Median (IQR) | 35 (30-42) | 50 (46-54) | 30 (24-38) | 38 (34-45) |
| **LGA** |  |  |  |  |
| Nguru | 6 | 6 | 6 | 7 |
| Karasuwa | 7 | 6 | 6 | 7 |

*FGD: Focus Group Discussion; IDI: In-Depth Interview; IQR: Interquartile Range; LGA: Local Government Area*

The following results present key facilitators and barriers influencing the acceptability and feasibility of measles vaccination and SQ-LNS distribution as part of the NutriVax-Measles trial across multiple levels. Findings are organized according to four main domains: (i) individual perceptions of vaccination, SQ-LNS, and the NutriVax strategy; (ii) social influence and community engagement, highlighting the role of community leaders, awareness campaigns, and male decision-makers; (iii) the quality and organization of health services supporting intervention delivery; and (iv) remaining structural and logistical barriers. This structure allows for a comprehensive understanding of how the intervention was perceived, implemented, and experienced in the field.

### Individual perceptions of measles vaccination, SQ-LNS and the NutriVax strategy

#### Vaccination

Most caregivers from both arms perceived vaccination as an effective means for preventing common vaccine-preventable diseases like measles and other childhood illnesses. Some also described that past experiences with outbreaks of measles and diphtheria, as well as fear of disease complications (e.g., blindness in measles) positively influenced their perceptions of immunization and served as a motivator for them to vaccinate their children.

> *“Because you see the children of your relatives and others who are vaccinated looking very healthy, so if you see that, you also get motivated to vaccinate your children**”- (FGD, Parents, Karasuwa LGA, NutriVax arm)***
>
> *“Because of what we have seen in the past (measles outbreak) […] we don’t wait, we take our children for vaccines.” - **(IDI, Parents, Karasuwa LGA, Control arm)***

However, in both arms, a few participants strongly held misconceptions that vaccines cause infertility. Others expressed concerns about post-immunization effects like fever, swelling, or children becoming irritable. To address this latest fear, collaboration with LGA and facility officials was strengthened to ensure a steady supply of paracetamol after vaccination in both arms.

> *“My worry is after a child gets vaccinated. …Last time, my baby had a fever and cried the whole night after the injection… [sighs] I don’t know if I can go through that again. But if there is a way to give fewer injections… or maybe a milder vaccine? because I am afraid of the side effects.”. **(IDI, Parents, Nguru LGA, NutriVax arm)***
>
> *“You will hear elders saying, why should we keep giving our children immunization when your parents gave you immunization, otherwise you would not have given birth. They say it prevents childbirth. Elders say it often.” **(IDI, Parents, Nguru LGA, NutriVax arm)***

#### SQ-LNS

SQ-LNS was also widely accepted by parents and caregivers in the NutriVax arm, who appreciated its perceived benefits for appetite, child health, and overall well-being of children after consistent use. Nearly all caregivers interviewed found the taste of SQ-LNS acceptable, and no HCWs reported any challenge related to it. Caregivers also found it easy to give to their children, either taken directly from the sachet or mixed with food.

> *“Since I started giving it, my child became more active and eats more food. I don’t stop because I want him to continue being healthy.”- **(FGD, Parents, Nguru LGA, NutriVax arm)***

A few mothers admitted to sharing the supplement with ineligible siblings within their family. Others declared to share their child ration outside their family under a system whereby each mother takes turns to distribute to other mothers who are not beneficiaries of the programme. Some SQ-LNS distributors also noted a few parents were borrowing children to obtain extra sachets. Yet, participants indicated that the small sachet design discouraged sales.

> *“The others [children] at home also want it, so I just give them small from the child’s own.” **- (IDI, Parents, Nguru LGA, NutriVax arm)***
>
> *“Because of its small size, some people don’t sell it. Leaving it small is better. If it becomes large, that is when they will sell it. (…) The small one is cheaper; it’s the bigger one they usually sell. Right now, the big one [Ready-to-use therapeutic food: RUTF] is sold for ₦450 per sachet, but this small one [SQ-LNS] is sold for ₦60 in the market.” **- (IDI, Parent, Nguru LGA, NutriVax arm)***

#### NutriVax strategy: combination of vaccination and SQ-LNS

Many caregivers from the NutriVax arm appreciated receiving SQ-LNS during the same day of vaccination sessions and explained that it gave them a tangible reason to maintain vaccination schedules. All interviewees from both community and health system indeed shared that the NutriVax strategy enhanced care-seeking behavior and acceptability of health facility visits, encouraging many parents who were previously hesitant to bring their children for immunization.

> *“When they gave the supplement, I saw that my child was eating well and became strong. That made me happy to continue coming for vaccination because I knew both helped my child.” **- (IDI, Parents, Nguru LGA, NutriVax arm)***
>
> *“Yes, it’s now easier to take children to the facility because this support [SQ-LNS] being given motivates them to go. In the past, you will find a child and their mother lying at home … the child is sick, no money for transport, no money for treatment …and they would say they would not go to the hospital. But now that this is being provided, it has greatly improved their children’s health.” - **(IDI, Community Health Workers, Nguru LGA, NutriVax Arm)***

### Community engagement, social influence, and gender norms influencing decision-making

As planned in the standard operational procedures, CNMs and volunteers organized several routine health education during immunization days in both arms as well as on session days when SQ-LNS was distributed in NutriVax arms. All interviewees agreed that such community mobilization was a key to reinforce awareness on vaccination and infant and young child feeding good practices in both arms, and on SQ-LNS in NutriVax arm, and encouraged caregivers to access healthcare facilities.

> *“Because of this organization [ALIMA], there are people from their team who go house to house, sensitizing community members and encouraging them to go to the hospital and explaining the benefits of doing so. Seeing this has truly increased the number of people going to the hospital now compared to the past.” - **(IDI, Community representative, Nguru LGA, Control Arm)***

Community leaders also brought their children first for vaccination, which helped build trust and enhance the acceptability of measles vaccination within the community in both arms. Some caregivers further explained that sharing success stories or attending healthcare facilities in groups motivated others to participate in the NutriVax programme.

> *“The traditional leader even starts by gathering his own children and grandchildren to get them vaccinated first. If there were any problem, it would happen to his children first. That is why it has been done not just once or twice, and no such problems have been seen. So even if someone tries to spread rumors, people don’t believe them**.” - (IDI, Community representative, Karasuwa LGA, Control Arm)***

In addition to the social influence and community engagement, gender norms were central in shaping care-seeking behavior in both trial arms. In the majority of families, fathers were the ones who made decisions regarding family care and provided money to cover transportation expenses. While most mothers required spousal approval before taking a child to the health center for vaccination, some men had negative perceptions regarding vaccines at the beginning of the trial. In response to that barrier, CNMs, religious leaders, and community representatives targeted and sensitized men at places where they gathered to socialize, markets where they conducted business, and mosques where they prayed in congregation. Several caregivers explained that fathers were thus more supportive, while some even accompanied their wives to facilities.

> *“Honestly, in the past, men were against taking children for vaccinations. If a woman tried, it could even cause serious issues in the marriage. But now, after repeated sensitization, they [men] understand the benefits of vaccines and will even be the ones to remind their wives that it’s time to take the child.” - **(FGD**, **Parent, Nguru LGA, NutriVax arm)***

Yet, some households still practiced traditional decision-making patterns at the end of the trial, as reported by a HCW.

> *“Some fathers do not accept immunization at all, no matter what you tell them. They are hard to convince. But mothers are usually more willing. The main problem is when fathers refuse to give transport money.” - **(FGD, Health Care Worker, Karasuwa LGA, NutriVax arm)***

### Quality of care and organization of health services

In both arms, participants shared increased attendance in healthcare facilities following sensitization activities, with a more pronounced increase following the introduction of SQ-LNS in the NutriVax arm. The resulting increase in attendance initially created challenges related to workload, waiting times, and service organization. Operational adjustments implemented over the course of the trial to reduce waiting time and improve the comfort of waiting areas were appreciated by stakeholders in both arms. Ultimately, participants’ accounts from the intervention arm indicated that co-delivery of vaccination and SQ-LNS was operationally feasible after adaptations were made to the organization of health services.

> *“It [workload] has really increased, because in the past, we used to wait for the women to come but now every Monday, they come in their numbers for measles vaccination… before now, we closed at the latest by two o’clock, but now we reach up to three or even five in the evening because more women come for the NutriVax [SQ-LNS].” - **(IDI, Health Worker, Karasuwa LGA, NutriVax arm)***
>
> *“We start [working at the clinic] early now. Mothers who come from far are the first to be attended to so they can go back before the sun becomes hot. It helps a lot because before, people complained of waiting for too long.”- **(IDI, Health Worker, Karasuwa LGA, Control arm)***
>
> *“Honestly, the introduction of the new register has been very useful. Previously, with the ration card, you would see a woman claim she lost it or say she couldn’t find it, but now with the register, it has really helped. Once people come, we just check the register… no delay. ”- **(IDI, Community Health Worker, Nguru LGA, NutriVax arm)***
>
> *“Previously, you would see mothers who brought their young children waiting under the sun because there was no tree or shade where it [SQ-LNS] was distributed. (…) now they collect it in comfort, with care, and on time, unlike before when it was noisy and overcrowded.”- **(IDI, Community Representative, Nguru LGA, NutriVaxarm)***

Interviewees reported no interruptions in vaccine availability in either arm, or in SQ-LNS in the NutriVax arm.

> *“We have not had a shortage of vaccines. Even when we need something, we call them and they bring it. The cold box is working, and the vaccines are always available.” - **(FGD, Health Worker, Karasuwa LGA, Control arm)***

Most caregivers in both arms also expressed satisfaction with the quality of care provided in health facilities, particularly regarding respectful attitude and courteous interactions of health workers, who took time to explain procedures and reassure them. While a few parents reported negative attitudes from some HCWs working in clinics in both arms and SQ-LNS distributors, measures implemented over the course of the trial to improve interpersonal-skills helped restore caregivers’ confidence and willingness to return for services.

> *“The attitude of the health workers is part of what makes people come more often. […] This reassurance makes people confident to return.“ **- (IDI, Community Representative, Karasuwa LGA, NutriVax arm)***

### Structural and logistical accessibility: persistent but mitigated barriers

In both arms, some caregivers, especially those in distant settlements still faced challenges related to poor road conditions, seasonal flooding, or heat. During the rainy season, roads to facilities often became impassable, forcing caregivers to postpone visits. Additionally, caregivers coming from distant settlements struggled with transportation costs, and they found it difficult to reach health facilities, particularly in emergency situations. Nevertheless, several interviewees reported that some parents, even if not all, manage to go to the healthcare centres for their children’s health, especially in the intervention arm where SQ-LNS was provided for free.

> *“In rural areas, distance and poverty are major issues. Some parents may want to bring their sick child but have nothing to eat themselves, let alone money for transport. They just sit and watch the child because they cannot afford to take them to the hospital.” - **(FGD, Community representative, Nguru LGA, NutriVax arm)***
>
> *“You know, with the current economic situation in Nigeria, many people don’t have money. Some even trek long distances to reach the hospital. As I mentioned earlier, some walk up to 16km just to get to the facility, especially mothers who come to receive the NutriVax [SQ-LNS] and also bring their sick children.” **- (IDI, Parents, Karasuwa LGA, NutriVax arm)***

Some caregivers also shared having maternal workload including farming or market that could prevent them from going to the healthcare facilities on time.

> *“Household chores and farm work keep me busy, during the rainy season. …and by the time I’m done, it’s mostly too late to go to the health center.” **- (IDI, Parent, Nguru LGA, Control arm)***

The provision of two- to three-month SQ-LNS rations for caregivers from remote areas, introduced during implementation to reduce the frequency of healthcare facility visits, was appreciated by parents.

> *“For those who come from distant villages, we give them two- or three-months’ ration at once so that they do not have to come every month. That has really helped because transport is costly for them.” **- (IDI, Community Health Worker, Karasuwa LGA, NutriVax arm)***

## Discussion

Overall, our findings showed that combining SQ-LNS distribution with routine measles immunization services was acceptable to caregivers, communities, and health-system stakeholders, and feasible to implement within primary healthcare services. However, successful implementation of both measles vaccination and SQ-LNS required adaptations to address barriers operating at individual, social, health-system, and structural levels (Figure 3).

**Figure 3.**
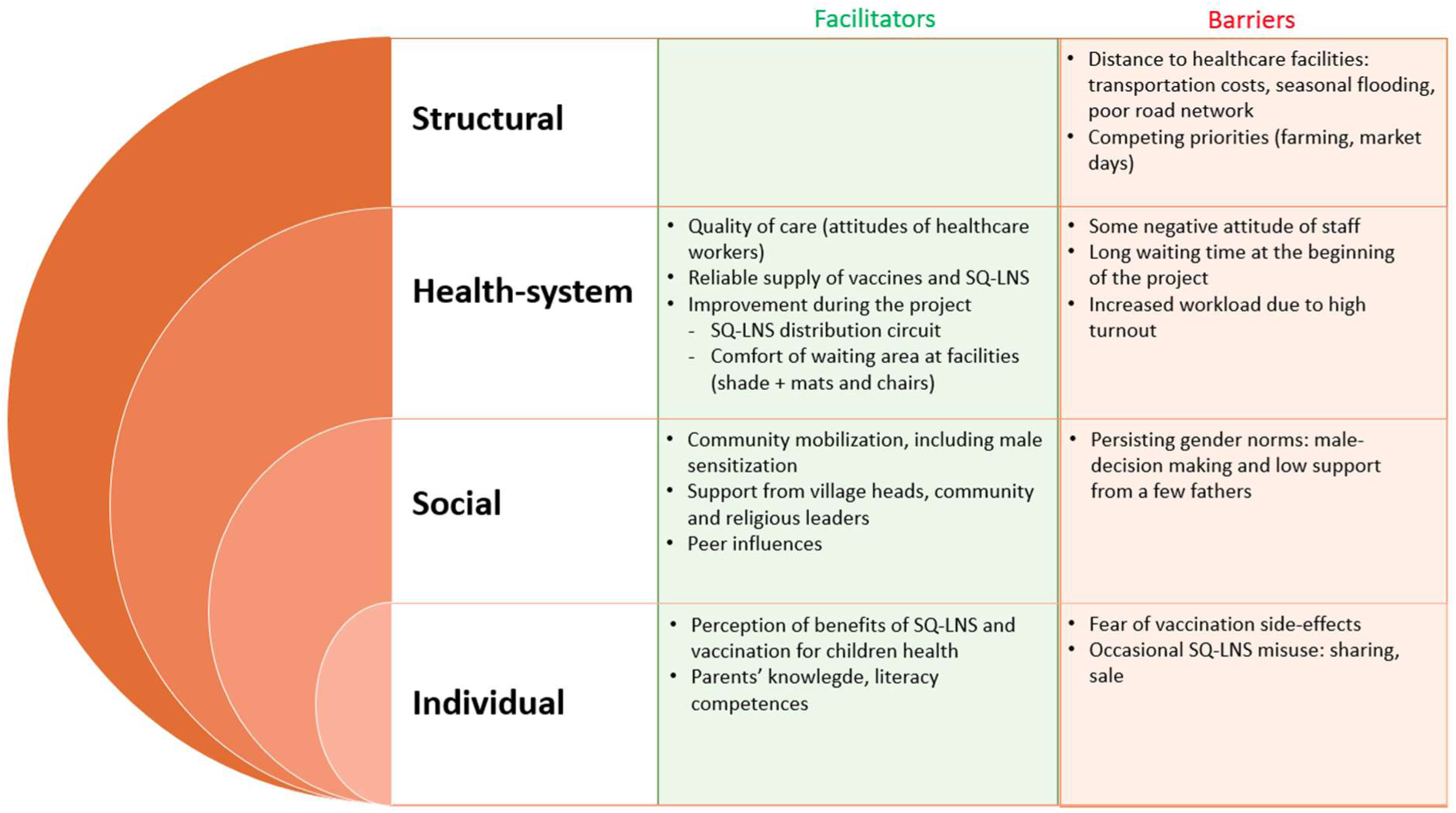
Summary of facilitators and barriers of implementing measles vaccination and SQ-LNS in Northern Nigeria.

Caregivers in the NutriVax arm perceived SQ-LNS as improving children’s appetite, activity, and health, in line with previous studies (28,29). These perceived benefits were reported as important motivations for attending health facilities, thereby creating additional opportunities for vaccination. This finding complements the quantitative results of the main NutriVax-Measles trial, which demonstrated a significant increase in measles vaccination uptake associated with the intervention (17). SQ-LNS was also valued by caregivers as a child health intervention in its own right. Some deviations from intended SQ-LNS use were reported, including sharing with siblings and other non-beneficiaries; however, the small sachet size may have limited this dynamic as well as its possible resale value, which highlights how design attributes can shape programme implementation and adherence (30).

At the end of the trial, most of the caregivers from both NutriVax and control arms perceived measles vaccination as an effective means of preventing severe childhood illness, with previous outbreak experience shaping positive attitudes towards immunization, as has been shown elsewhere (31,32). While fear of adverse events following immunization, as well as widespread misconceptions about vaccines, such as beliefs that they cause infertility, could lead to vaccine hesitancy (33–35), community mobilization was consistently described by participants as an important facilitator of vaccination uptake across both trial arms. The engagement of traditional and religious leaders, together with CNMs, contributed to shifting some erroneous beliefs and enhancing public confidence towards vaccination, consistent with previous work (13,36,37). Some of the community leaders even modeled positive health behavior for their communities by having their children vaccinated first, highlighting the influence of social networks and trusted individuals on vaccination behaviors (38,39). These qualitative findings complement the quantitative findings showing increased measles vaccination uptake between baseline and endline in both trial arms (17), and reinforce evidence that community engagement is a critical component of vaccination strategies in low-coverage settings.

Gender norms remained a key determinant of care-seeking. Many mothers reported requiring spousal approval before bringing their children to be vaccinated. Similar findings have been documented across African settings (40), including evidence from Ghana showing higher vaccination coverage when men were engaged in health-related decisions (41). In the NutriVax trial, initial concerns among fathers were addressed through targeted sensitization activities conducted in social and religious gathering spaces, following which participants reported increased male support for immunization. These findings emphasize that improving childhood vaccination coverage requires engaging not only mothers but also other household decision-makers for the long-term success of child health interventions in northern Nigeria and similar settings.

From a health-system perspective, the findings suggest that combining SQ-LNS distribution with routine immunization was feasible, but that feasibility was supported in part by iterative adaptations made during implementation rather than being an inherent feature of the original intervention package. Increased caregiver attendance initially resulted in longer waiting times in PHCCs were SQ-LNS was provided, and higher workloads for healthcare workers. In response, adaptations were introduced to improve registration systems, patient flow and waiting areas, and these changes were positively perceived by both healthcare workers and caregivers. These findings highlight the importance of adaptive implementation when increased demand may place additional pressure on existing health-system capacity. While SQ-LNS was distributed only in PHCCs within the NutriVax-Measles trial, the decentralization within smaller health facilities would likely have helped balance the additional workload, and to some extent, alleviate overcrowding at PHCCs. Co-delivery of nutrition and immunization services should therefore not be viewed simply as the addition of a new service, but as an intervention that may require ongoing adjustment and strategic investment in service organization to respond to emerging implementation challenges. This interpretation is consistent with previous literature suggesting that service-level and organizational interventions can improve patient experience and healthcare delivery outcomes (42). The attitudes of HCWs were also cited as important in influencing caregivers’ experiences and willingness to use services, as previously described (43). In this context, HCWs and SQ-LNS distributors were specifically trained to improve interpersonal skills and make sure they have a respectful attitude. Caregivers subsequently expressed high satisfaction with the quality of interactions at health facilities. Such capacity-building approaches may require repeated reinforcement to support sustained changes in provider behavior over time.

Persistent structural, geographical, and logistical barriers, including long distances between home and health facilities as well as competing household priorities, also limited equitable access to vaccination and SQ-LNS, as also found in the quantitative findings (17). Distance to health facilities, poor road conditions, seasonal flooding, and transportation costs remained important challenges, especially since many caregivers lived in remote settlements and had limited economic resources. These findings are consistent with previous studies identifying geographical and financial constraints as major barriers to vaccination uptake in African settings (40,44–46). In addition, competing livelihood and household priorities, including farming activities and market days, limited caregivers’ availability to attend health facilities, particularly during peak farming periods. These opportunity costs represent important determinants of healthcare utilization (12,47,48), and demonstrate that positive perceptions of an intervention alone are insufficient to overcome broader structural constraints. Several adaptations were implemented in PHCCs where SQ-LNS was distributed, to mitigate these barriers, including providing two- to three-month SQ-LNS rations (instead of a one-month ration) for caregivers living far from health facilities, as was anticipated in the study protocol, as well as prioritizing them in the queue during visits. Although these adaptations could not fully eliminate access constraints, they reduced the frequency of required visits and improved the feasibility of participation for vulnerable households. Future scale-up strategies should consider such context-specific adaptations to ensure equitable access. The results also highlight the importance of decentralizing the distribution of SQ-LNS to smaller health facilities, where immunization activities are routinely conducted, to improve accessibility.

This study adds evidence on the acceptability and feasibility of combining measles vaccination with SQ-LNS distribution in West Africa. The inclusion of stakeholders from both community and health-system levels provided a comprehensive understanding of the NutriVax intervention implementation, and the convergence of findings across these stakeholder groups strengthens the validity of the results. Data collection by local qualitative research assistants fluent in local languages likely facilitated trust, reduced interpretation bias and enabled a more contextually grounded understanding of participants’ perceptions and experiences. The use of established conceptual frameworks further supported a systematic interpretation of barriers and facilitators to measles vaccination and SQ-LNS implementation.

Yet, this study has several limitations. First, the qualitative analysis was mainly based on a rapid thematic approach using interview summaries rather than systematic coding of full transcripts. This approach was selected to allow timely feedback during implementation but may have reduced analytical granularity and limited the identification of more nuanced perspectives in participants’ narratives; nevertheless, the transcripts made it possible to retrieve the most relevant quotes following the rapid analysis to illustrate the different findings. Second, FGDs were conducted only among participants from intervention wards, limiting direct comparison of group-level perspectives between intervention and control arms. In addition, social desirability bias cannot be excluded, although the consistency of findings across different stakeholder groups provides some reassurance regarding the robustness of the major themes.

## Conclusion

This study showed that co-delivering preventive SQ-LNS with routine immunizations was acceptable and feasible at primary healthcare settings in an insecure, resource-constrained context in Northern Nigeria that faces persistent high malnutrition rates and low vaccination coverage. The perceived benefits for children’s health, along with the convenience of receiving both services in a single visit, contributed to the intervention’s acceptability within the community. The findings also highlight the importance of adapting intervention delivery to address implementation barriers. Future scale-up of such integrated care models in settings facing a similar dual threat to child survival will require sustained investment in community mobilization, including targeted sensitization efforts focused on men; health-system capacity-building; and structural efforts to decentralize the provision of SQ-LNS alongside immunization services, bringing them as close as possible beneficiaries to improve equitable access to care.

## Data Availability

A summary matrix with de-identified data will be made available subject to certain conditions within 3 months after publication through the IRD DataSuds repository (https://dataverse.ird.fr/) with an assigned study DOI, in accordance with institutional requirements for the data curation process. The code will remain available for at least 15 years. Requests for accessing the code should be submitted through the DataSuds repository using the DOI and will be reviewed by the corresponding author at. A decision will be provided within 4 weeks of receiving the request. Approved applicants will receive access to the dataset and code through the IRD DataSuds repository.

## Author contributions

Conceptualization: MP, CC, KPQP

Data curation: SIA, SAK, AMS, AGG, AG

Formal analysis: SIA, MP

Funding acquisition: RB, CC, KPQP

Project administration: AMA, IA, ECU, SB, NSM, BWG, UC, HMA

Supervision: MP

Validation: JN, CC, KPQP

Writing - original draft: SIA, JN, MP

Writing - review and editing: all co-authors

## Funding

This study was funded by the Eleanor Crook Foundation (ECF) and Gavi, the Vaccine Alliance. The Alliance for International Medical Action (ALIMA), University of Bordeaux and the Yobe State Ministry of Health of Yobe State, Nigeria, provided operational support for project implementation. The funders had no role in the study design, data collection, analysis, or interpretation of this study.

## Acknowledgments

We wish to thank the ALIMA teams in Yobe and Abuja for their support and the individuals who participated in this study. The authors also acknowledge the support and fruitful collaboration of the Yobe State MoH in implementing this study including their initiatives to ensure stakeholder engagement at the level of the State, Nguru and Karasuwa Local Government Areas, and at the community levels.

## Competing interests

KP reports having served on the Social Purposes Advisory Committee of Nutriset, a major producer of lipid-based nutritional supplements. The remaining authors report no conflicts of interest in this work.

## Supplementary files

**Supplementary file 1.** SRQR guidelines

**Supplementary file 2.** Distribution of population (0-24 Months) and health facility coverage across qualitative evaluation study wards

**Supplementary file 3.** Interview guides

**Supplementary file 4.** Summary grid completed for each of the interviews

**Supplementary file 5.** Analysis grids for each theme based on the conceptual frameworks

## Notes

### Clinical Trial

NCT06387511

### Author Declarations

Ethical approval was obtained from the Yobe State Ministry of Health and Human Services Health Research Ethics Committee (YOHREC) on February 14th, 2024 (MOH/GEN/747_February 2024) and from the Ethics Evaluation Committee of the French National Institute for Health and in France from the National Institute for Health and Medical Research (Inserm) Ethics Evaluation Committee (24-1091-20240402/IRB0000388). Written informed consent was obtained from all participants before the interview. Participants were informed about the purpose of the study and their right to withdraw at any time without any consequences. Recordings were stored on a secure drive accessible only to the qualitative research team.

