## Supplementary File 1 for "Acceptability and feasibility of measles vaccination, small-quantity lipid-based nutrient supplementation, and their co-delivery among children aged 6–23 months in northern Nigeria: a qualitative study"

The SRQR reporting checklist

For checking that qualitative health research articles can be understood and used by everyone

**Manuscript:** Acceptability and feasibility of combining Small-Quantity Lipid-based Nutrient Supplements (SQ-LNS) distributions with routine immunization among children 6-23 months of age in Northeastern Nigeria: a qualitative study

|  | Item Description | Location (or reason for not reporting) |
| --- | --- | --- |
| **Title & Abstract** |  |  |
| [Title](https:/resources.equator-network.org/reporting-guidelines/srqr/items/title.html?utm_source=srqr&utm_medium=checklist&utm_campaign=1_1) | Describe the nature and topic of the study. Identify the study as qualitative or indicate the approach or data collection methods. | Title, p.1 |
| [Abstract](https:/resources.equator-network.org/reporting-guidelines/srqr/items/abstract.html?utm_source=srqr&utm_medium=checklist&utm_campaign=1_1) | Summarise the key elements of the study using the abstract format of the intended publication. | Abstract – Methods, p.3 |
| **Introduction** |  |  |
| [Problem Formulation](https:/resources.equator-network.org/reporting-guidelines/srqr/items/problem-formulation.html?utm_source=srqr&utm_medium=checklist&utm_campaign=1_1) | Describe the problem/phenomenon studied, its significance, relevant theory and empirical work, and gaps in current knowledge. | Introduction, p.5, l.122-145 |
| [Purpose or research question](https:/resources.equator-network.org/reporting-guidelines/srqr/items/purpose.html?utm_source=srqr&utm_medium=checklist&utm_campaign=1_1) | Describe the purpose of the study and specific objectives or questions. | Introduction, p.5, l.149-156 |
| **Methods** |  |  |
| [Qualitative approach and research paradigm](https:/resources.equator-network.org/reporting-guidelines/srqr/items/qualitative-approach.html?utm_source=srqr&utm_medium=checklist&utm_campaign=1_1) | Describe your qualitative approach, your guiding theory (if appropriate), and research paradigm, and reasons for your choices. | Methods – Study design, p.7, l.161-164  & Conceptual framework, p.9, l.217-226 |
| [Researcher characteristics and reflexivity](https:/resources.equator-network.org/reporting-guidelines/srqr/items/researcher-characteristics-and-reflexivity.html?utm_source=srqr&utm_medium=checklist&utm_campaign=1_1) | Describe how researchers’ characteristics may influence the research, including personal attributes, qualifications/experience, relationship with participants, assumptions, and/or presuppositions; potential or actual interaction between researchers’ characteristics and the research questions, approach, methods, results and/or transferability. | Methods – Data collection p.9, l.232-234  & Discussion – p.21, l.584-586 |
| [Context](https:/resources.equator-network.org/reporting-guidelines/srqr/items/context.html?utm_source=srqr&utm_medium=checklist&utm_campaign=1_1) | Describe the setting/site(s) in which the study was conducted, why it was selected, and any other salient contextual factors that may influence the study. | Methods – Study setting, p.6-7 |
| [Sampling strategy](https:/resources.equator-network.org/reporting-guidelines/srqr/items/sampling-strategy.html?utm_source=srqr&utm_medium=checklist&utm_campaign=1_1) | Describe how and why research participants, documents, or events were selected; criteria for deciding when no further sampling was necessary, and the rationale for those criteria. | Methods – Study population and sampling strategy, p.7-8  & Table 1, p.8-9 |
| [Ethical issues pertaining to human subjects](https:/resources.equator-network.org/reporting-guidelines/srqr/items/ethics.html?utm_source=srqr&utm_medium=checklist&utm_campaign=1_1) | Describe any approval by an appropriate ethics review board and participant consent, or explain any lack thereof. Describe any other confidentiality and data security issues. | Methods – Ethics, p.10-11 |
| [Data collection methods](https:/resources.equator-network.org/reporting-guidelines/srqr/items/data-collection-methods.html?utm_source=srqr&utm_medium=checklist&utm_campaign=1_1) | Describe the types of data collected; details of data collection procedures including (as appropriate) start and stop dates of data collection and analysis, iterative process, triangulation of sources/methods, and modification of procedures in response to evolving study findings. Describe your rationale for these choices. | Methods – Data collection, p.9 |
| [Data collection instruments and technologies](https:/resources.equator-network.org/reporting-guidelines/srqr/items/data-collection-instruments.html?utm_source=srqr&utm_medium=checklist&utm_campaign=1_1) | Describe any instruments (e.g., interview guides, questionnaires) and devices (e.g., audio recorders) used for data collection; describe if/how the instrument(s) changed over the course of the study. | Methods – Data collection, p.9 |
| [Units of study](https:/resources.equator-network.org/reporting-guidelines/srqr/items/units-of-study.html?utm_source=srqr&utm_medium=checklist&utm_campaign=1_1) | Describe the number and relevant characteristics of participants, documents, or events included in the study. Describe the level of participation. | Methods – Table 1, p.8-9  & Results – Table 2, p.12 |
| [Data processing](https:/resources.equator-network.org/reporting-guidelines/srqr/items/data-processing.html?utm_source=srqr&utm_medium=checklist&utm_campaign=1_1) | Describe the methods for processing data prior to and during analysis, including transcription, data entry, data management and security, verification of data integrity, data coding, and anonymisation / deidentification of excerpts. | Methods – Data analysis, p.9-10  & Methods – Ethics, p.11 |
| [Data analysis](https:/resources.equator-network.org/reporting-guidelines/srqr/items/data-analysis.html?utm_source=srqr&utm_medium=checklist&utm_campaign=1_1) | Describe the process by which inferences, themes, etc. were identified and developed, including the researchers involved in data analysis; usually references a specific paradigm or approach. Describe why you chose this process. | Methods – Data analysis, p.9-10 |
| [Techniques to enhance trustworthiness](https:/resources.equator-network.org/reporting-guidelines/srqr/items/trustworthiness.html?utm_source=srqr&utm_medium=checklist&utm_campaign=1_1) | Describe any techniques to enhance trustworthiness and credibility of data analysis,(e.g., member checking, triangulation, audit trail). Describe why you chose these techniques. | Methods – Data analysis, p.10 |
| **Results** |  |  |
| [Synthesis and interpretation](https:/resources.equator-network.org/reporting-guidelines/srqr/items/synthesis-and-interpretation.html?utm_source=srqr&utm_medium=checklist&utm_campaign=1_1) | Describe the main findings (e.g., interpretations, inferences, and themes); might include development of a theory or model, or integration with prior research or theory. | Results, p.13-18 |
| [Links to empirical data](https:/resources.equator-network.org/reporting-guidelines/srqr/items/links-to-empirical-data.html?utm_source=srqr&utm_medium=checklist&utm_campaign=1_1) | Provide evidence (e.g., quotes, field notes, text excerpts, photographs) to substantiate analytic findings. | Results, p.13-18 |
| **Discussion** |  |  |
| [Integration with prior work, implications, transferability, and contribution(s) to the field](https:/resources.equator-network.org/reporting-guidelines/srqr/items/integration-with-prior-work.html?utm_source=srqr&utm_medium=checklist&utm_campaign=1_1) | Summarize the main findings, explain how findings and conclusions connect to, support, elaborate on, or challenge conclusions of earlier scholarship; discuss the scope of application/generalizability; identify unique contribution(s) to scholarship in a discipline or field. | Discussion, p.18-22 |
| [Limitations](https:/resources.equator-network.org/reporting-guidelines/srqr/items/limitations.html?utm_source=srqr&utm_medium=checklist&utm_campaign=1_1) | Discuss the trustworthiness and limitations of findings | Discussion, p.21-22, l.579-594 |
| **Other** |  |  |
| [Conflicts of interest](https:/resources.equator-network.org/reporting-guidelines/srqr/items/conflicts-of-interest.html?utm_source=srqr&utm_medium=checklist&utm_campaign=1_1) | Describe any potential sources of influence or perceived influence on study conduct and conclusions. Describe how these were managed. | Competing interest, p.2 |
| [Funding](https:/resources.equator-network.org/reporting-guidelines/srqr/items/funding.html?utm_source=srqr&utm_medium=checklist&utm_campaign=1_1) | Describe sources of funding and other support. Describe the role of funders in data collection, interpretation, and reporting. | Funding, p.2 |
