## Supplementary File 2 for "Acceptability and feasibility of measles vaccination, small-quantity lipid-based nutrient supplementation, and their co-delivery among children aged 6–23 months in northern Nigeria: a qualitative study"

**Supplementary file 1.** Distribution of population (0-24 Months) and health facility coverage across qualitative evaluation study wards

| **LGA** | **Ward** | **Population size** | **Distribution of 0-24 per PHCC/PHC/HP** |
| --- | --- | --- | --- |
| **Nguru** | Afunori (I) | 21532 | 1 PHCC, 2 PHCs, 1 HP |
| **Nguru** | Dabule (I) | 15730 | 1 PHC, 2 HPs |
| **Nguru** | Maja Kura (I) | 23650 | 1 PHCC, 1 PHC, 2 HPs |
| **Nguru** | Bulabulin (C) | 25988 | 1 PHCC, 2 PHCs, 1 HP |
| **Nguru** | Ngarbi (C) | 25039 | 1 PHC, 2 HPs |
| **Nguru** | Tsohon Nguru (C) | 31723 | 1 PHCC, 2 PHCs |
| **Karasuwa** | Fajiganari (I) | 15934 | 1 PHC, 2 HPs |
| **Karasuwa** | Karasuwa-Garin Guna (I) | 17211 | 1 PHCC, 2 PHCs |
| **Karasuwa** | Wachakal (I) | 16445 | 1 PHC, 2 HPs |
| **Karasuwa** | Jajimaji (C) | 28982 | 1 PHCC, 3 PHCs, 1 HP |
| **Karasuwa** | Jajeri (C) | 23840 | 1 PHC, 2 HPs |
| **Karasuwa** | Waro (C) | 22678 | 1 PHC, 2 HPs |

*PHCC: Primary Health Care Centre; PHC: Primary Health Care; HP: Health Post*

*I: Intervention arm; C: Control arm*
