## Supplementary File 3 for "Acceptability and feasibility of measles vaccination, small-quantity lipid-based nutrient supplementation, and their co-delivery among children aged 6–23 months in northern Nigeria: a qualitative study"

**NutriVax – Measles**

**Interview guides**

### Interview Guide - Parents/Guardian

#### Socio -demographic characteristics

| **Gender** | **□** Female □ Male |
| --- | --- |
| **Age** | …… years old |
| **Educational Level** | □ None □ Primary □ Secondary □ Others : ………… |
| **Relationship to Baby** | □ Mother □ Father □ Others : ………… |
| **Marital Status** | □ Married □ Separated □ Widowed |
| **Age at Marriage** |  |
| **Number of Children** |  |
| **Household composition** |  |
| **Household income** | □ <NGN 50,000 □ NGN 50 -100,000 □ >NGN 100,000 |
| **Have access to** | □ HP □ PHC □ PHCC |

- If you don't understand certain questions and need me to rephrase them, please don't hesitate to let me know.
- Also, please note that there are no right or wrong answers to the questions I will ask you.
- Before we begin, do you have any questions about this interview?
- Do you agree to let me record the interview?

#### Theme 1. Presentation

**1. Child’s status**

###### If I am right, you have a baby under two years old?

###### Age

###### Sex

###### Health status

#### Theme 2. Access to health care

**2a. Experiences and Logistical Challenges**

###### What are your experiences of going to the health center for your baby?

###### *Main medical reason to access health center*

###### Have you experienced any challenges accessing the health centre?

###### *Please explain.*

###### *Explore especially transportation (distance, means of transport) and financial challenges (e.g., missed work, costs) to determine how this affects their ability to access care.*

##### 2b. Decision-making

###### Who makes decisions in the caregiver’s household regarding the children’s health?

###### Can you describe a recent situation where a decision was made about your child’s health?

###### *Including taking them to the vaccine clinics?*

###### *Differences according, to the sex of the baby?*

###### What do you think regarding fathers' involvement in seeking healthcare?

###### *Probe on vaccinations, provision of transport.*

- *Are there any cultural or social factors influencing decision-making around vaccinations/seeking healthcare services?*

##### 2c. Quality of Care

###### What is your perception of the quality of care in the health facility?

###### *Probe on opening hours, waiting times and availability of healthcare workers during your visits?*

###### *How does these factors impact on your trust in the facility?*

###### Does the profile of healthcare workers influence your interaction with healthcare workers?

###### *Probe on Sex and Age, years of experience*

###### *If yes, how?*

#### Theme 3. Vaccination

##### 3a. Knowledge and Perception

###### What do you know about vaccination among children?

###### How have you been informed about vaccination?

###### *Probe on community sensitizations*

###### What do you think about vaccination among children?

###### How do you perceive the importance of vaccines in preventing illnesses in children above 6 months?

###### *Probe on measles vaccination*

###### Can you share your experience with health education on vaccination during antenatal visits?

###### Have you ever heard any rumours or misconceptions about vaccination in your community?

###### *If* ***yes probe:*** *mention the misconceptions, where did she hear them from?*

##### 3b. Vaccination Status and Decision-making

###### Has your child had any vaccines? Which ones?

###### *Probe on measles vaccination: number of doses? place of vaccination?*

###### What are the main reasons that have influenced your decision to vaccinate (or not vaccinate) your child?

###### *Explore facilitators/motivations and barriers/challenges like:*

###### ***Logistical:*** *costs, transport, living environment, hours of opening*

###### ***Perceptions/knowledge:*** *community awareness, healthcare worker advice, HCW interactions*

###### ***Community support / norms:*** *community mobilizers, caregiver support, common practices*

###### ***In intervention arms:*** *SQ-LNS availability*

###### *Explore specific barriers/facilitators to Measles Vaccination (compared to other vaccines)?*

###### Does the decision to vaccinate your child differ according to the sex of the baby? Why?

###### Are there any specific challenges or barriers that made it difficult for you to get your child vaccinated in subsequent visits?

###### *Is the vaccination available in the same health centre you used to go?*

##### 3c. Community Influences

###### What are common beliefs and behaviours about child vaccinations in your community?

###### How do these beliefs and behaviours influence your decisions?

##### 3d. Experience of vaccination

###### Thinking about vaccination day for your child, could you tell me what happens? What you liked and what you didn’t like?

###### *Before you leave home: what did you need to do to prepare, how did you travel to the facility, with who*

###### *At the* *vaccination place: what happened in the waiting area or queue*

###### *When it was your child’s turn to get vaccine: what did healthcare workers talk to you about vaccine and explain the vaccine and its benefice? how did you feel when you talked to them? and when your child received its first vaccination?*

###### *After you child has had the vaccine: how did you travel, what happened at home*

##### 3e. Potential additional Motivators

###### If your child is not vaccinated, what would motivate you to go to the health center for vaccination?

###### *Explore if it differs between parents? And by the sex of the baby?*

###### Would you be more likely to vaccinate your child if you received additional support or service?

###### *Which one?*

###### *In control arm: such as SQ-LNS?*

#### Theme 4. SQ-LNS supplementation *[in the NutriVax arm only]*

##### 4a. General Awareness and Perception

###### What have you heard about SQ-LNS supplementation?

###### How have you been informed about SQ-LNS?

###### *Probe on community sensitizations: Community Nutrition mobilizers, other caregivers, community members, interactions with health workers?*

###### How does your community view SQ-LNS supplementation?

##### 4b. Usage and Adherence

###### Has your baby already taken SQ-LNS?

###### *Why or why not?*

###### What do you like about SQ-LNS?

###### *Probe on benefits of SQ-LNS*

###### And what you don’t like?

###### If your baby has already taken SQ-LNS, could you explain how it goes regarding giving SQ-LNS daily to your child?

###### *Explore factors like: taste, complexity of uptake, child’s health status, living conditions, priorities, common practices, and method of uptake (sachet or mixed with food)*

###### *Are their days you missed giving SQ-LNS to your baby?*

###### Are there challenges in using SQ-LNS exclusively for your child? / Do you know if the SQ-LNS has been shared with other children?

###### *In which contexts?*

- How do you ensure your child takes SQ-LNS daily?
  - *Are there any specific routines or reminders?*

##### 4c. Experience and perception regarding the Distribution of SQ-LNS

###### Thinking about the day of SQ-LNS collection for your child, could you tell me what happens? What you liked and what you didn’t like?

###### *Before you leave home: what did you need to do to prepare, how did you travel to the facility, with who?*

###### *What was your experience like at the distribution site, especially in the waiting area or queue?*

###### *When it was your child’s turn: what did the healthcare workers talk to you about SQ-LNS, and how did you feel during the interaction?*

###### More generally, what do you think about the way in which SQ-LNS is distributed?

###### *Use of ration card, Number of rations, Waiting time*

###### Have you observed any change regarding waiting areas in the facility you used to go?

###### *Probe on comfort (mats, benches, chairs, tables)*

###### *Waiting time*

###### Have you noticed any other changes in how SQ-LNS is given out in the last three months compared to before?

#### Theme 5. Sustainability

*In the Control arm: explain the NutriVax strategy: in some villages/healthcare facilities, children receive a nutritional supplementation called SQ-LNS when their vaccination status is reviewed.*

##### 5a. Perceptions regarding overall access to vaccination

###### In your opinion, how could it be possible to improve the access to vaccination in your community?

###### What types of support or incentives would encourage families in your community to vaccinate their children ?

###### (e.g., reminders, free transport, SQ-LNS, community support)

###### What do you think about community mobilization that have been implemented in your community to improve access to vaccination?

###### *Including community mobilization focused on men?*

*In the Control arm: explain the NutriVax strategy: in some villages/healthcare facilities, children receive a nutritional supplementation called SQ-LNS when their vaccination status is reviewed.*

##### 5b. Perceptions regarding the NutriVax strategy and sustainability

###### What do you think about offering SQ-LNS to motivate parents/guardians to vaccinate their children?

###### Perception of sustainability

- *In intervention arm: Do you think that this intervention should continue in your community?*
- *In control arm: Do you think that this intervention should be implemented in your community? Why?*
- *If yes, do you see any conditions for it to be well implemented? How could the NutriVax strategy be adapted/scaled up to improve the access to vaccination in your community?*

### Interview Guide – Community representatives

#### Socio -demographic characteristics

| **Gender** | **□** Female □ Male |
| --- | --- |
| **Age** | …… years old |
| **Educational Level** | □ None □ Primary □ Secondary □ Others : ………… |
| **Role within the Community** |  |
| **Duration in Leadership Role** |  |

- If you don't understand certain questions and need me to rephrase them, please don't hesitate to let me know.
- Also, please note that there are no right or wrong answers to the questions I will ask you.
- Before we begin, do you have any questions about this interview ?
- Do you agree to let me record the interview ?

#### Theme 1. Role and understanding of the project

##### 1a. Community Role

###### Could you explain your role within the community?

##### 1b. Awareness of NutriVax Project

###### What do you know about the NutriVax project?

###### How was the project introduced to your community, and what was the initial reaction?

##### 1c. Community Influence

###### How does your role influence health-related initiatives in the community, such as vaccination programs? Give any example.

###### *In particular as part of the NutriVax project?*

- How do you think your role could be utilized more effectively in promoting vaccination programs?

#### Theme 2. Access to health care centres

##### 2a. General Perceptions

###### According to what you observe in your community, what would you say about access to care for babies under two years old?

###### *What are the main medical reasons families go to clinic?*

###### *Differences by sex of the babies?*

###### What are the main barriers families face in accessing care?

###### *Explore factors like :*

###### ***Logistical/financial:*** *distance to health centres, transportation, costs, missed work*

###### ***Quality of care:*** *clinic operating hours, waiting times, availability of healthcare workers, quality of care*

###### Explore facilitators

##### 2b. Decision-making

###### In this community, who makes decisions in the caregiver’s household regarding their children’s health?

###### *Including taking them to the vaccine clinics?*

###### *Differences according to the sex of the baby?*

###### Are there differences in how fathers and mothers bring children to the clinic?

###### *Are mothers able to go by themselves to the health centre when their children are sick / needs vaccination?*

###### Do cultural or community beliefs influence decisions about seeking healthcare for babies?

###### *Does that include vaccination?*

##### 2c. Quality of Care and Trust in Services

###### [if not explored before] How do the parents in your community perceive the quality of care in the health facilities?

###### *Probe on opening hours, waiting times and availability of healthcare workers during your visits?*

###### Do you think parents trust the health services provided at the clinics?

###### *Why or why not?*

###### *Does it differs according to sex or age of healthcare workers?*

###### How do you think the community’s perception of healthcare services could be improved?

##### 2d. Community Events

###### Have there been any events within your district that have disrupted access to healthcare for babies in the last three months?

###### *Examples: flooding, insecurity, rumors*

###### Probe on facilitators

#### Theme 3. Vaccination

##### 3a. Perception of Vaccination

###### What do you think about vaccination among children?

###### *Probe on measles vaccination*

##### 3b. Parental Reactions

###### What do people in your community think about measles vaccination?

###### *Explore differences in reactions based on the age, gender and parents' backgrounds*

###### *Explore differences according to the sex and age of the baby*

###### In your opinion, what motivates members of your community to vaccinate their children in general?

###### What are the main barriers?

###### *Explore facilitators/motivations and barriers/challenges like:*

###### ***Financial/Logistical:*** *costs, transport, living environment, hours of opening*

###### ***Perceptions/knowledge:*** *rumor, community awareness, healthcare worker advice, HCW interactions*

###### ***Community support / norms:*** *caregiver support, common practices, gender differences*

###### ***SQ-LNS*** *availability*

###### Are you able to overcome these challenges? How?

###### Are there any specific barriers regarding measles vaccination (compared to other vaccines)? And facilitators?

###### *Have there been any specific beliefs or rumours in the community that affect how families view measles vaccination?*

##### 3c. Community Events

###### Have there been any events within your district that have disrupted access to vaccination for babies?

###### Probe on facilitators

#### Theme 4. SQ-LNS supplementation *[in the NutriVax arm only]*

##### 4a. Perception and Awareness

###### What do you think about the SQ-LNS supplementation?

###### *How do you think SQ-LNS could benefit children in your community?*

- How do you think SQ-LNS could be integrated into existing community health programs or activities?

##### 4b. Community Reactions

###### According to what you have observed in your community, how do people perceive SQ-LNS?

###### How is SQ-LNS given/administered daily to eligible children?

###### *Daily uptake? Why yes? Why not?*

###### *Sharing practices? Sale of SQ-LNS? If yes in which context?*

##### 4c. External context

###### Have there been any events within your district that have facilitated or disrupted SQ-LNS supplementation?

###### *Including community mobilization regarding SQ-LNS use?*

#### Theme 5. Relations with NutriVax project team

##### 5a. General interactions

- What are your interactions with people working for the NutriVax project?
- Have you played a specific role in the implementation of the NutriVax project?

###### If yes

###### What has your role involved?

###### How did you find your experience in this role?

- *What challenges have you experienced?*
- *What successes have you experienced?*

#### Theme 6. Sustainability

##### 6a. Perceptions regarding overall access to vaccination

###### In your opinion, how could it be possible to improve the access to vaccination in your community?

###### What types of support or incentives would encourage families in your community to vaccinate their children ?

###### (e.g., reminders, free transport, SQ-LNS, community support)

###### What do you think about community mobilization that have been implemented in your community to improve access to vaccination?

###### *Including community mobilization focused on men?*

*In the Control arm: explain the NutriVax strategy: in some villages/healthcare facilities, children receive a nutritional supplementation called SQ-LNS when their vaccination status is reviewed.*

##### 6b. Perceptions regarding the NutriVax strategy and sustainability

###### What do you think about offering SQ-LNS as an incentive to motivate parents/guardians to vaccinate their children?

- *W*o*uld offering SQ-LNS motivates parents to vaccinate their child? Why?*

###### Perception of sustainability

- *In intervention arm: Do you think that this intervention should continue in your community?*
- *In control arm: Do you think that this intervention should be implemented in your community?*

###### If yes, do you see any conditions for it to be well implemented? How could the NutriVax strategy be adapted/scaled up to improve the access to vaccination in your community?

### Interview Guide – Community health workers

#### Socio -demographic characteristics

| **Gender** | **□** Female □ Male |
| --- | --- |
| **Age** | …… years old |
| **Educational Level** | □ Certificate □ Diploma □Degree Other: ………… |
| **Years of experience** |  |
| **Access to PHCC/PHC/HP** | □ HP □ PHC □ PHCC |

- If you don't understand certain questions and need me to rephrase them, please don't hesitate to let me know.
- Also, please note that there are no right or wrong answers to the questions I will ask you.
- Before we begin, do you have any questions about this interview ?
- Do you agree to let me record the interview ?

#### Theme 1. Role and understanding of the project

##### 1a. Role and Motivation to take part in the NutriVax projet

###### Could you explain me what is your role in this NutriVax project?

###### Could you tell me what motivates you to participate in this project?

###### What challenges do you face in your role, and how do you overcome them?

#### Theme 2. Access to health care centres

##### 2a. General Perceptions

###### According to what you observe, what would you say about access to care for babies under two years old in your community?

###### *What are the main reasons families go to clinic?*

###### *Differences by sex of the babies?*

###### What are the main barriers families face in accessing care?

###### *Explore factors like:*

###### ***Logistical/financial:*** *distance to health centres, transportation, costs, missed work*

###### ***Quality of care:*** *clinic operating hours, waiting times, availability of healthcare workers, quality of care*

###### Explore facilitators

##### 2b. Decision-making

###### In this community, who makes decisions in the caregiver’s household regarding their children’s health?

###### *Including taking them to the vaccine clinics?*

###### *Differences according to the sex of the baby?*

###### Are there differences in how fathers and mothers bring children to the clinic?

###### *Are mothers able to go by themselves to the health centre when their children are sick / needs vaccination?*

###### Do cultural or community beliefs influence decisions about seeking healthcare for babies?

###### *Does that include vaccination?*

##### 2c. Quality of Care and Trust in Services

###### [if not explored before] How do the parents in your community perceive the quality of care in the health facilities?

###### *Probe on opening hours, waiting times and availability of healthcare workers during your visits?*

###### Do you think parents trust the health services provided at the clinics?

###### *Why or why not?*

###### *Does it differs according to sex or age of healthcare workers?*

##### 2d. Community Events

###### Have there been any events within your district that have disrupted access to healthcare for babies?

###### *Examples: flooding, insecurity, rumors*

###### Probe on facilitators

#### Theme 3. Vaccination

##### 3a. Perception of Vaccination

###### What do you think about vaccination among children?

###### *Probe on measles vaccination*

##### 3b. Parental Reactions

###### What are your experiences when you talk to parents/guardians about vaccination?

###### *Explore differences in reactions based on the age, gender and parents' backgrounds*

###### *Explore differences according to the sex and age of the baby*

###### In your opinion, what motivates members of your community to vaccinate their children in general?

###### What are the main barriers?

###### *Explore facilitators/motivations and barriers/challenges like:*

###### ***Financial/Logistical:*** *costs, transport, living environment, hours of opening*

###### ***Perceptions/knowledge:*** *rumor, community awareness, healthcare worker advice, HCW interactions*

###### ***Community support / norms:*** *caregiver support, common practices, gender differences*

###### ***SQ-LNS*** *availability*

###### Are you able to overcome these challenges? How?

###### Are there any specific barriers regarding measles vaccination (compared to other vaccines)? And facilitators?

###### *Have there been any specific beliefs or rumours in the community that affect how families view measles vaccination?*

###### *If yes, how have you addressed them?*

##### 3e. Community Events

###### Have there been any events within your district that have disrupted access to vaccination for babies?

###### Probe on facilitators

#### Theme 4. SQ-LNS supplementation *[in the NutriVax arm only]*

##### 4a. Perception and Awareness

###### What do you think about the SQ-LNS supplementation?

###### *How do you think SQ-LNS could benefit children in your community?*

##### 4b. Community Reactions

###### What are the reactions of the members of your community when you talk to them about SQ-LNS?

###### Have you encountered any misconceptions or skepticism about SQ-LNS?

###### If so, how have you addressed them?

##### 4c. Adherence and Practices

###### What do you think about the adherence to the SQ-LNS among children in your community?

###### Based on what you observe or what parents/guardians share with you?

###### *Explore Factors like: taste, complexity of uptake, exchange for money, child’s health, priorities, common practices, and method of uptake (sachet or mixed with food).*

###### What factors influence whether parents continue using SQ-LNS consistently over time?

##### 4d. Sharing Practices

###### Have you already observe/be aware of SQL-NS sharing between children or with other families in your community?

###### *In which contexts?*

###### Have you already observe/be aware of SQL-NS sale in your community?

###### *In which contexts?*

###### Are there cultural practices that might influence how SQ-LNS is shared or distributed among children?

##### 4e. External context

###### Have there been any events within your district that have facilitated or disrupted SQ-LNS supplementation?

###### *Including community mobilization regarding SQ-LNS use?*

#### Theme 5. Relations with NutriVax project team

5a. General Perception

- How do functional and logistical issues affect the activities of NutriVax services in your facility?

###### *Explore potential shortages of SQ_LNS/vaccines, lack of staff, sub-optimal organization, long queue etc.*

- How the implementation of NutriVax in the HP/PHC/PHCC has changed your daily activities?
  - *What are the positive effects? The negative effects?*
  - *Has your workload changed since joining the NutriVax project?*
    - *If so, how do you balance your tasks?*

###### How does the NutriVax intervention fit into the tasks you were doing before?

##### 5b. Collaboration and Dynamics

###### What are your interactions with other people working for the NutriVax project?

###### What challenges or successes have you experienced in collaborating with the NutriVax team?

#### Theme 6. Sustainability

##### 6a. Perceptions regarding overall access to vaccination

###### In your opinion, how could it be possible to improve the access to vaccination in your community?

###### What types of support or incentives would encourage families in your community to vaccinate their children ?

###### (e.g., reminders, free transport, SQ-LNS, community support)

###### What do you think about community mobilization that have been implemented in your community to improve access to vaccination?

###### *Including community mobilization focused on men?*

*In the Control arm: explain the NutriVax strategy: in some villages/healthcare facilities, children receive a nutritional supplementation called SQ-LNS when their vaccination status is reviewed.*

##### 6b. Perceptions regarding the NutriVax strategy and sustainability

###### What do you think about offering SQ-LNS as an incentive to motivate parents/guardians to vaccinate their children?

- *W*o*uld offering SQ-LNS motivates parents to vaccinate their child? Why?*

###### Perception of sustainability

- *In intervention arm: Do you think that this intervention should continue in your community?*
- *In control arm: Do you think that this intervention should be implemented in your community?*

###### If yes, do you see any conditions for it to be well implemented? How could the NutriVax strategy be adapted/scaled up to improve the access to vaccination in your community?

### Interview Guide – Health care workers

#### Socio -demographic characteristics

| **Gender** | **□** Female □ Male |
| --- | --- |
| **Age** |  |
| **Educational Level** | □ Certificate □ Diploma □ Degree □ Others: ………… |
| **Role** |  |
| **Years of experience** |  |
| **Type of Health Structure** | □ HP □ PHC □ PHCC |

- If you don't understand certain questions and need me to rephrase them, please don't hesitate to let me know.
- Also, please note that there are no right or wrong answers to the questions I will ask you.
- Before we begin, do you have any questions about this interview ?
- Do you agree to let me record the interview ?

#### Theme 1. Role and understanding of the project

##### 1a. Role and Motivation to take part in the NutriVax projet

###### Could you explain me what is your role in this NutriVax project?

###### Could you tell me what motivates you to participate in this project?

###### What challenges do you face in your role, and how do you overcome them?

#### Theme 2. Access to health care centres

##### 2a. General Perceptions

###### What would you say about access to care for babies under two years old in this community?

###### *What are the main reasons families go to clinic?*

###### *Differences by sex of the babies?*

###### Besides vaccination, have you observed any changes in the use of other services since the NutriVax project started?

###### *Explain in detail any changes*

###### What are the main barriers families face in accessing care?

###### *Explore factors like :*

###### ***Logistical/financial****: distance to health centres, transportation, costs, missed work*

###### ***Quality of care****: clinic operating hours, waiting times, availability of healthcare workers, quality of care*

###### Explore facilitators

##### 2b. Decision-making

###### In this community, who makes decisions in the caregiver’s household regarding their children’s health?

###### *Including taking them to the vaccine clinics?*

###### *Differences according to the sex of the baby?*

###### Are there differences in how fathers and mothers bring children to the clinic?

###### *Are mothers able to go by themselves to the health centre when their children are sick / needs vaccination?*

###### Do cultural or community beliefs influence decisions about seeking healthcare for babies?

###### *Does that include vaccination?*

##### 2c. Community Events and Adaptations

###### Have there been any events within your district that have disrupted access to healthcare for babies?

###### *Examples: flooding, insecurity, rumors*

###### Probe on facilitators

###### What do you think regarding waiting areas in your facility?

###### *Probe on comfort (mats, benches, chairs, tables)*

###### *Changes over time*

#### Theme 3. Vaccination

3a. General Perception

###### Can you summarize the procedure of immunization, starting once a family arrives at the centres for vaccination?

###### What do you think about vaccination activities among children in your health facility?

###### *What is working well? What challenges are you facing?*

###### *Explore potential shortages of vaccines, cold chain, packaging of vaccines, lack of staff, sub-optimal organization, long queue etc.*

###### Are there specifics barriers that specifically affect measles vaccination uptake at facility level?

###### *Explore differences between the first and second doses of measles vaccines*

##### 3b. Parental Reactions

What is the reaction of parents/guardians? when you offer them vaccination ?

###### *Explore differences in reactions based on the age, gender and parents' backgrounds*

###### *Explore differences according to the sex and age of the baby.*

###### What factors do parents consider most when deciding whether to vaccinate their children?

###### In your opinion, what motivates members of your community to vaccinate their children?

###### And what are the main barriers/challenges?

###### *Explore facilitators/motivations and barriers/challenges like:*

###### ***Logistical:*** *costs, transport, living environment, hours of opening*

###### ***Perceptions/knowledge:*** *rumor, community awareness, healthcare worker advice, HCW interactions*

###### ***Community support / norms:*** *caregiver support, common practices, gender differences including sex of child*

###### ***SQ-LNS*** *availability*

###### Are you able to overcome these challenges? How?

###### *What helps those who are hesitant about getting their children vaccinated?*

###### Are there any specific barriers regarding measles vaccination (compared to other vaccines)? And facilitators?

###### *Have there been any specific beliefs or rumours in the community that affect how families view measles vaccination?*

###### *If yes, how have you addressed them?*

##### 3c. Community Events

###### Have there been any events within your district that have disrupted access to vaccination for babies?

###### Probe on facilitators

#### Theme 4. SQ-LNS supplementation *[in the NutriVax arm only]*

##### 4a. Perceptions and Awareness

###### What do you think about the SQ-LNS supplementation?

###### *How do you think SQ-LNS could benefit children in your community?*

##### 4b. Community Reactions

###### What are the reactions of the members of your community when you talk to them about SQ-LNS?

###### Have you encountered any misconceptions or skepticism about SQ-LNS?

###### If so, how have you addressed them?

##### 4c. Adherence and Practices

###### What do you think about the adherence to the SQ-LNS among children in your community?

###### Based on what you observe or what parents/guardians share with you?

###### *Explore Factors like: taste, complexity of uptake, exchange for money, child’s health, priorities, common practices, and method of uptake (sachet or mixed with food).*

###### What factors influence whether parents continue using SQ-LNS consistently over time?

##### 4d. Sharing Practices

###### Have you already observe/be aware of SQL-NS sharing between children or with other families in your community?

###### *In which contexts?*

###### Have you already observe/be aware of SQL-NS sale in your community?

###### *In which contexts?*

###### Are there cultural practices that might influence how SQ-LNS is shared or distributed among children?

##### 4f. External context

###### Have there been any events within your district that have facilitated or disrupted SQ-LNS supplementation?

###### *Including community mobilization regarding SQ-LNS use?*

#### Theme 5. Working conditions and relations with NutriVax project team

5a. General Perception

###### How do functional and logistical issues affect the activities of NutriVax services in your facility?

###### *Explore potential shortages of SQ_LNS/vaccines, lack of staff, sub-optimal organization, long queue etc.*

###### How the implementation of NutriVax in the HP/PHC/PHCC has changed your daily activities?

*Explore: vaccination, SQ-LNS*

###### *What are the positive effects? The negative effects?*

- *Has your workload changed since joining the NutriVax project?*
  - *If so, how do you balance your tasks?*

###### How does the NutriVax intervention fit into the tasks you were doing before?

##### 5b. Collaboration and Dynamics

###### What are your interactions with other people working for the NutriVax project?

###### What challenges or successes have you experienced in collaborating with the NutriVax team?

#### Theme 6. Sustainability

##### 6a. Perceptions regarding overall access to vaccination

###### In your opinion, how could it be possible to improve the access to vaccination in your community?

###### What types of support or incentives would encourage families in your community to vaccinate their children ?

###### (e.g., reminders, free transport, SQ-LNS, community support)

###### What do you think about community mobilization that have been implemented in your community to improve access to vaccination?

###### *Including community mobilization focused on men?*

*In the Control arm: explain the NutriVax strategy: in some villages/healthcare facilities, children receive a nutritional supplementation called SQ-LNS when their vaccination status is reviewed.*

##### 6b. Perceptions regarding the NutriVax strategy and sustainability

###### What do you think about offering SQ-LNS as an incentive to motivate parents/guardians to vaccinate their children?

- *W*o*uld offering SQ-LNS motivates parents to vaccinate their child? Why?*

###### Perception of sustainability

- *In intervention arm: Do you think that this intervention should continue in your community?*
- *In control arm: Do you think that this intervention should be implemented in your community?*

###### If yes, do you see any conditions for it to be well implemented? How could the NutriVax strategy be adapted/scaled up to improve the access to vaccination in your community?

### Interview Guide - FGD Parents/Guardian

#### Socio -demographic characteristics

***Complete this table for each participant of the FGD***

| **Gender** | **□** Female □ Male |
| --- | --- |
| **Age** | …… years old |
| **Educational Level** | □ None □ Primary □ Secondary □ Others : ………… |
| **Relationship to Baby** | □ Mother □ Father □ Others : ………… |
| **Marital Status** | □ Married □ Separated □ Widowed |
| **Age at Marriage** |  |
| **Number of Children** |  |
| **Household composition** |  |
| **Household income** | □ <NGN 50,000 □ NGN 50 -100,000 □ >NGN 100,000 |
| **Have access to** | □ HP □ PHC □ PHCC |

**Introduction – reminders**

- If you don't understand certain questions and need me to rephrase them, please don't hesitate to let me know.
- Also, please note that there are no right or wrong answers to the questions I will ask you.
- Before we begin, do you have any questions about this interview?
- Do you agree to let me record the interview?

#### Theme 1. Access to health care

**1a. You are parents/guardians of a baby aged <2 years old. Could you tell me what are you experiences of going to the health centre for your baby?**

###### What are the main medical reasons to access healthcare centres for your baby?

###### Have you experienced any challenges accessing healthcare centres? (logistical, financial, other…)

###### What is your perception of the quality of care in healthcare centres? (competences of HCW, waiting time, quality of services…)

###### Are there common practices or beliefs that support or discourage these visits?

##### 1b. In you families/households, how decisions are made regarding the children’s health, including on vaccination?

###### Who makes the decisions?

###### Does the decision differ according to the sex of the baby?

###### Are there any cultural or social factors influencing decision-making around vaccinations/seeking healthcare services?

###### How do you think it influence children’s health?

#### Theme 2. Vaccination

##### 2a. What do you know about vaccination among children?

###### Especially regarding measles vaccination

###### Can you share your experience with health education on vaccination?

###### *During antenatal visits*

###### *Through community sensitizations*

##### 2b. What do you think about vaccination for your child?

###### Has your child had any vaccines? Where was your child vaccinated?

###### What are the main reasons that have influenced your decision to vaccinate (or not vaccinate) your child?

###### *Explore facilitators/motivations and barriers/challenges like:*

###### ***Logistical:*** *costs, transport, living environment, hours of opening*

###### ***Perceptions/knowledge:*** *community awareness, healthcare worker advice, HCW interactions*

###### ***Community support / norms:*** *community mobilizers, caregiver support, common practices, person who make the decision regarding vaccination*

###### ***In intervention arms:*** *SQ-LNS availability*

###### *Explore specific barriers/facilitators to Measles Vaccination (compared to other vaccines)?*

###### What are common beliefs about vaccinations in your community?

###### How do these beliefs influence your decisions?

#### Theme 3. SQ-LNS supplementation

##### 3a. What do you think about SQ-LNS supplementation in general?

###### What are the benefits you know?

##### 3b. What do you think about the way in which SQ-LNS is distributed? Have you noticed any changes in the last three months compared to before?

###### Use of ration card

###### Number of rations

###### Waiting time

###### Waiting areas (mats, benches, chairs, tables)

##### 3c. Could you share your experience of giving (or not) SQ-LNS to your baby?

###### Why giving or not giving?

###### What do you like about SQ-LNS? And what you don’t like?

###### If your baby has already taken SQ-LNS, could you explain how it goes regarding giving SQ-LNS daily to your child?

###### *Explore Factors like: taste, complexity of uptake, child’s health status, living conditions, priorities, common practices, and method of uptake (sachet or mixed with food)*

###### *Are their days you missed giving SQ-LNS to your baby?*

###### Are there challenges in using SQ-LNS exclusively for your child? / Do you know if the SQ-LNS has been shared with other children, or used differently?

###### *In which contexts?*

- How do you ensure your child takes SQ-LNS daily?
  - *Are there any specific routines or reminders?*

##### 3d. Have you noticed or heard of SQ-LNS being sold in the community? *If so, where or by whom?*

##### How common do you think this practice is?

##### Why do you think some people choose to sell SQ-LNS instead of giving it to their children?

###### *Probe on poverty, or lack of understanding about its purpose?*

##### What do you think could be done to discourage the sale of SQ-LNS in the community?

###### *Probe on education or monitoring?*

#### Theme 4. Sustainability of the NutriVax intervention

It is almost the end of the NutriVax-measles trial.

##### 4a. Perceptions regarding overall access to vaccination

###### In your opinion, how could it be possible to improve the access to vaccination in your community after the trial ends?

###### What types of support or incentives would encourage families in your community to vaccinate their children ?

###### (e.g., reminders, free transport, SQ-LNS, community support)

###### What do you think about community mobilization that have been implemented in your community to improve access to vaccination?

###### *Including community mobilization focused on men?*

##### 4b. Perceptions regarding the NutriVax strategy and sustainability

###### What do you think about offering SQ-LNS as an incentive to motivate parents/guardians to vaccinate their children?

- *W*o*uld offering SQ-LNS motivates parents to vaccinate their child? Why?*

###### Perception of sustainability: Do you think that this intervention should continue in your community? And extended in your country? Why?

###### If yes, do you see any conditions for it to be well implemented? How could the NutriVax strategy be adapted/scaled up to improve the access to vaccination in your community?

### Interview Guide – FGD Community representatives

#### Socio -demographic characteristics

***Complete this table for each participant of the FGD***

| **Gender** | **□** Female □ Male |
| --- | --- |
| **Age** | …… years old |
| **Educational Level** | □ None □ Primary □ Secondary □ Others : ………… |
| **Role within the Community** |  |
| **Duration in Leadership Role** |  |

**Introduction – reminders**

- If you don't understand certain questions and need me to rephrase them, please don't hesitate to let me know.
- Also, please note that there are no right or wrong answers to the questions I will ask you.
- Before we begin, do you have any questions about this interview?
- Do you agree to let me record the interview?

#### Theme 1. Access to health care centres

**We will first begin with general questions regarding access to healthcare centres in you communities.**

##### 1a. What would you say about access to care for babies <2 years old in general?

###### What are the main medical reasons families go to clinic?

###### Are there differences by sex of the babies?

###### What are the main barriers families face in accessing care?

###### *Explore factors like :*

###### ***Logistical/financial:*** *distance to health centres, transportation, costs, missed work*

###### ***Quality of care:*** *clinic operating hours, waiting times, availability of healthcare workers, quality of care*

###### Explore facilitators

###### Do cultural or community beliefs influence decisions about seeking healthcare for babies?

##### 1b. In your community, how decisions are made regarding the children’s health, including on vaccination?

###### Who makes the decisions? Are mothers able to go by themselves to the health centre when their children are sick / needs vaccination?

###### Are there differences in how fathers and mothers bring children to the clinic?

###### Does the decision differ according to the sex of the baby?

###### Are there any cultural or social factors influencing decision-making around vaccinations/seeking healthcare services?

##### 1c. Have there been any events within your district/community that have disrupted access to healthcare for babies?

###### Probe on facilitators

#### Theme 2. Vaccination

##### 2a. What do you think about vaccination among children in general?

##### 2b. Can you share your experience with health education on vaccination in your community?

###### *Probe on community sensitizations*

##### 2c. What do you see in your community regarding children vaccination?

###### What motivates members to vaccinate their children?

###### What are the main barriers?

###### *Explore facilitators/motivations and barriers/challenges like:*

###### ***Financial/Logistical:*** *costs, transport, living environment, hours of opening*

###### ***Perceptions/knowledge:*** *rumor, community awareness, healthcare worker advice, HCW interactions*

###### ***Community support / norms:*** *caregiver support, common practices, gender differences, person who make the decision regarding vaccination*

###### ***SQ-LNS*** *availability*

###### Are there any specific barriers regarding measles vaccination (compared to other vaccines)? facilitators? (beliefs, rumours, pain…)

##### 2d. Have there been any events within your district/community that have disrupted access to vaccination for babies?

###### Probe on facilitators

#### Theme 3. SQ-LNS supplementation

##### 3a. What do you think about the SQ-LNS supplementation?

###### How do you think SQ-LNS could benefit children in your community?

###### What do you think about offering SQ-LNS as an incentive to motivate parents/guardians to vaccinate their children?

##### 3b. According to what you have observed in your community, how do people perceive SQ-LNS and give it to their children?

###### What are the challenges of using SQ-LNS daily? Or facilitators?

###### *Explore Factors like: taste, complexity of uptake, child’s health status, living conditions, priorities, common practices, and method of uptake (sachet or mixed with food)*

###### Have you already observe/be aware of SQL-NS sharing between children, or used differently, in your community? If yes, in which context?

##### 3c. Have you noticed or heard of SQ-LNS being sold in the community? *If so, where or by whom?*

##### How common do you think this practice is?

##### Why do you think some people choose to sell SQ-LNS instead of giving it to their children?

###### *Probe on poverty, or lack of understanding about its purpose?*

##### What do you think could be done to discourage the sale of SQ-LNS in the community?

###### *Probe on education or monitoring?*

##### 3d. What do you think about how SQ-LNS is offered/distributed in your community? Have you noticed any changes in the last three months compared to before?

###### Use of ration card

###### Number of rations

###### Waiting time

###### Waiting areas (mats, benches, chairs, tables)

##### 3e. Have there been any events within your district/community that have facilitated or disrupted SQ-LNS supplementation?

#### Theme 4. Roles and relations with NutriVax project team

##### 5a. Has there been any evolution regarding your involvement in the NutriVax project since the beginning of the project?

###### What was your role?

###### How did you find your experience in this role? (challenges, successes…)

##### 5b. How do you perceived your relations with others people working for the NutriVax project?

#### Theme 5. Sustainability of the NutriVax intervention

It is almost the end of the NutriVax-measles trial.

##### 5a. Perceptions regarding overall access to vaccination

###### In your opinion, how could it be possible to improve the access to vaccination in your community after the trial ends?

###### What types of support or incentives would encourage families in your community to vaccinate their children ?

###### (e.g., reminders, free transport, SQ-LNS, community support)

###### What do you think about community mobilization that have been implemented in your community to improve access to vaccination?

###### *Including community mobilization focused on men?*

##### 5b. Perceptions regarding the NutriVax strategy and sustainability

###### What do you think about offering SQ-LNS as an incentive to motivate parents/guardians to vaccinate their children?

- *W*o*uld offering SQ-LNS motivates parents to vaccinate their child? Why?*

###### Perception of sustainability: Do you think that this intervention should continue in your community? And extended in your country? Why?

###### If yes, do you see any conditions for it to be well implemented? How could the NutriVax strategy be adapted/scaled up to improve the access to vaccination in your community?

### Interview Guide – FGD Community health workers

#### Socio -demographic characteristics

***Complete this table for each participant of the FGD***

| **Gender** | **□** Female □ Male |
| --- | --- |
| **Age** | …… years old |
| **Educational Level** | □ Certificate □ Diploma □Degree Other: ………… |
| **Years of experience** |  |
| **Access to PHCC/PHC/HP** | □ HP □ PHC □ PHCC |

**Introduction – reminders**

- If you don't understand certain questions and need me to rephrase them, please don't hesitate to let me know.
- Also, please note that there are no right or wrong answers to the questions I will ask you.
- Before we begin, do you have any questions about this interview?
- Do you agree to let me record the interview?

#### Theme 1. Access to health care centres

**We will first begin with general questions regarding access to healthcare centres in you communities.**

##### 1a. What would you say about access to care for babies <2 years old in general?

###### What are the main medical reasons families go to clinic?

###### Are there differences by sex of the babies?

###### What are the main barriers families face in accessing care?

###### *Explore factors like :*

###### ***Logistical/financial:*** *distance to health centres, transportation, costs, missed work*

###### ***Quality of care:*** *clinic operating hours, waiting times, availability of healthcare workers, quality of care*

###### Explore facilitators

###### Do cultural or community beliefs influence decisions about seeking healthcare for babies?

##### 1b. In your community, who makes decisions in the caregiver’s household regarding their children’s health?

###### Who makes the decisions? Are mothers able to go by themselves to the health centre when their children are sick / needs vaccination?

###### Are there differences in how fathers and mothers bring children to the clinic?

###### Does the decision differ according to the sex of the baby?

###### Are there any cultural or social factors influencing decision-making around vaccinations/seeking healthcare services?

##### 1c. Have there been any events within your district/community that have disrupted access to healthcare for babies?

###### Probe on facilitators

#### Theme 2. Vaccination

##### 2a. What do you think about vaccination among children in general?

##### 2b. Can you share your experience with health education on vaccination in your community?

###### *Probe on community sensitizations*

##### 2c. What do you see in your community regarding children vaccination?

###### What are your experiences when you talk to parents/guardians about vaccination?

###### *Explore differences in reactions based on the* ***age, gender and parents' backgrounds***

###### *Explore differences according to the sex and age of the baby*

###### What motivates members to vaccinate their children?

###### What are the main barriers?

###### *Explore facilitators/motivations and barriers/challenges like:*

###### ***Financial/Logistical:*** *costs, transport, living environment, hours of opening*

###### ***Perceptions/knowledge:*** *rumor, community awareness, healthcare worker advice, HCW interactions*

###### ***Community support / norms:*** *caregiver support, common practices, gender differences, person who make the decision regarding vaccination*

###### ***SQ-LNS*** *availability*

###### Are there any specific barriers regarding measles vaccination (compared to other vaccines)? facilitators? (beliefs, rumours, pain…)

###### Which strategies do you use to motivate parents/guardians to accept vaccination?

##### 3d. Have there been any events within your district/community that have disrupted access to vaccination for babies?

###### Probe on facilitators

#### Theme 4. SQ-LNS supplementation

##### 4a. What do you think about the SQ-LNS supplementation?

###### How do you think SQ-LNS could benefit children in your community?

###### What do you think about offering SQ-LNS as an incentive to motivate parents/guardians to vaccinate their children?

##### 4b. According to what you have observed in your community, how do people perceive SQ-LNS and give it to their children?

###### Have you encountered any misconceptions or skepticism about SQ-LNS?

###### If so, how have you addressed them?

###### What are the challenges of using SQ-LNS daily? Or facilitators?

###### *Explore Factors like: taste, complexity of uptake, child’s health status, living conditions, priorities, common practices, and method of uptake (sachet or mixed with food)*

###### Have you already observe/be aware of SQL-NS sharing between children, or used differently, in your community? If yes, in which context?

##### 4b. Have you noticed or heard about the sale of SQ-LNS in your community ?

- If so, where or by whom ?
- How common do you think this practice is ?
- Why do you think some people choose to sell SQ-LNS instead of giving their children ?

###### *Probe on poverty, or lack of understanding about its purpose?*

- Do you think selling SQ-LNS affects the program’s ability to improve child nutrition and vaccination rates ? How ?
- Does this create any challenges for your work as a CHW ?
- What do you think could be done to discourage the sale of SQ-LNS in the community ?
  - *Probe on education or monitoring could help ?*

##### 4d. What do you think about how SQ-LNS is offered/distributed in your community? Have you noticed any changes in the last three months compared to before?

###### Use of ration card

###### Number of rations

###### Waiting time

###### Waiting areas (mats, benches, chairs, tables)

##### 4e. Have there been any events within your district/community that have facilitated or disrupted SQ-LNS supplementation?

#### Theme 4. Roles and relations with NutriVax project team

##### 5a. Has there been any evolution regarding your involvement in the NutriVax project since the beginning of the project?

###### What was your role?

###### How did you find your experience in this role? (challenges, successes…)

- How the implementation of NutriVax in the HP/PHC/PHCC has changed your daily activities? (including workload)
  - *What are the positive effects? The negative effects? How do you balance your tasks?*

##### 5b. How do you perceived your relations with others people working for the NutriVax project?

##### 5c. How do you functional and logistical issues affect the activities of NutriVax services in your community/facility?

###### *Explore potential shortages of SQ_LNS/vaccines, lack of staff, sub-optimal organization, long queue etc.*

#### Theme 5. Sustainability of the NutriVax intervention

##### 5a. Perceptions regarding overall access to vaccination

###### In your opinion, how could it be possible to improve the access to vaccination in your community after the trial ends?

###### What types of support or incentives would encourage families in your community to vaccinate their children ?

###### (e.g., reminders, free transport, SQ-LNS, community support)

###### What do you think about community mobilization that have been implemented in your community to improve access to vaccination?

###### *Including community mobilization focused on men?*

##### 5b. Perceptions regarding the NutriVax strategy and sustainability

###### What do you think about offering SQ-LNS as an incentive to motivate parents/guardians to vaccinate their children?

- *W*o*uld offering SQ-LNS motivates parents to vaccinate their child? Why?*

###### Perception of sustainability: Do you think that this intervention should continue in your community? And extended in your country? Why?

###### If yes, do you see any conditions for it to be well implemented? How could the NutriVax strategy be adapted/scaled up to improve the access to vaccination in your community?

### Interview Guide – FGD Health care workers

#### Socio -demographic characteristics

***Complete this table for each participant of the FGD***

| **Gender** | **□** Female □ Male |
| --- | --- |
| **Age** |  |
| **Educational Level** | □ Certificate □ Diploma □ Degree □ Others: ………… |
| **Role** |  |
| **Years of experience** |  |
| **Type of Health Structure** | □ HP □ PHC □ PHCC |

**Introduction – reminders**

- If you don't understand certain questions and need me to rephrase them, please don't hesitate to let me know.
- Also, please note that there are no right or wrong answers to the questions I will ask you.
- Before we begin, do you have any questions about this interview?
- Do you agree to let me record the interview?

#### Theme 1. Access to health care centres

##### 1a. What would you say about access to care for babies <2 years old in general?

###### What are the main medical reasons families go to clinic?

###### Are there differences by sex of the babies?

###### What are the main barriers families face in accessing care?

###### *Explore factors like :*

###### ***Logistical/financial:*** *distance to health centres, transportation, costs, missed work*

###### ***Quality of care:*** *clinic operating hours, waiting times, availability of healthcare workers, quality of care*

###### Explore facilitators

###### Are there differences in how fathers and mothers bring children to the clinic?

##### 1b. Besides vaccination, have you observed any changes in the use of other services since the NutriVax project started?

##### 1c. Have there been any events within your district/community that have disrupted access to healthcare for babies?

###### Probe on facilitators

#### Theme 2. Vaccination

##### 2a. What are your experience about offering vaccination for children to parents/guardians?

###### *Explore differences in reactions based on the* ***age, gender and parents' backgrounds***

###### *Explore differences according to the sex and age of the baby*

###### What motivates members to vaccinate their children?

###### What are the main barriers?

###### *Explore facilitators/motivations and barriers/challenges like:*

###### ***Financial/Logistical:*** *costs, transport, living environment, hours of opening*

###### ***Perceptions/knowledge:*** *rumor, community awareness, healthcare worker advice, HCW interactions*

###### ***Community support / norms:*** *caregiver support, common practices, gender differences, person who make the decision regarding vaccination*

###### ***SQ-LNS*** *availability*

###### Are there any specific barriers regarding measles vaccination (compared to other vaccines)? And facilitators? (beliefs, rumours, pain…)

###### Which strategies do you use to motivate parents/guardians to accept vaccination?

##### 2b. Have you experienced any logistical challenges about offering vaccination to children in your healthcare facility? And what is going well?

###### *Explore potential shortages of vaccines, cold chain, packaging of vaccines, lack of staff, sub-optimal organization, long queue etc.*

##### 2c. Have there been any events within your district/community that have disrupted access to vaccination for babies?

###### Probe on facilitators

#### Theme 3. SQ-LNS supplementation

##### 3a. What do you think about the SQ-LNS supplementation?

###### How do you think SQ-LNS could benefit children in your community?

###### What do you think about offering SQ-LNS as an incentive to motivate parents/guardians to vaccinate their children?

##### 3b. According to what you have observed, how do people perceive SQ-LNS and give it to their children?

###### What are the reactions of the members of your community when you talk to them about SQ-LNS?

###### Have you encountered any misconceptions or skepticism about SQ-LNS?

###### If so, how have you addressed them?

###### What are the challenges of using SQ-LNS daily? Or facilitators?

###### *Explore Factors like: taste, complexity of uptake, child’s health status, living conditions, priorities, common practices, and method of uptake (sachet or mixed with food)*

###### Have you already observe/be aware of SQL-NS sharing between children, or used differently, in your community? If yes, in which context?

##### 3c. Have you noticed or heard about the sale of SQ-LNS in your community ?

- If so, where or by whom ?
- How common do you think this practice is ?
- Why do you think some people choose to sell SQ-LNS instead of giving their children ?

###### *Probe on poverty, or lack of understanding about its purpose?*

- Do you think selling SQ-LNS affects the program’s ability to improve child nutrition and vaccination rates ? How ?
- Does this create any challenges for your work as a CHW ?
- What do you think could be done to discourage the sale of SQ-LNS in the community ?
  - *Probe on education or monitoring could help ?*

##### 3d. Have there been any events within your district/community that have facilitated or disrupted SQ-LNS supplementation?

#### Theme 4. Roles and relations with NutriVax project team

##### 4a. Has there been any evolution regarding your involvement in the NutriVax project since the beginning of the project?

###### How did you find your experience in this role? (challenges, successes…)

- How the implementation of NutriVax in the HP/PHC/PHCC has changed your daily activities? (including workload)
  - *What are the positive effects? The negative effects? How do you balance your tasks?*

##### 4b. How do you perceived your relations with others people working for the NutriVax project?

##### 4c. How do you functional and logistical issues affect the activities of NutriVax services in your community/facility?

###### *Explore potential shortages of SQ_LNS/vaccines, lack of staff, sub-optimal organization, long queue etc.*

#### Theme 5. Sustainability of the NutriVax intervention

##### 5a. Perceptions regarding overall access to vaccination

###### In your opinion, how could it be possible to improve the access to vaccination in your community after the trial ends?

###### What types of support or incentives would encourage families in your community to vaccinate their children ?

###### (e.g., reminders, free transport, SQ-LNS, community support)

###### What do you think about community mobilization that have been implemented in your community to improve access to vaccination?

###### *Including community mobilization focused on men?*

##### 5b. Perceptions regarding the NutriVax strategy and sustainability

###### What do you think about offering SQ-LNS as an incentive to motivate parents/guardians to vaccinate their children?

- *W*o*uld offering SQ-LNS motivates parents to vaccinate their child? Why?*

###### Perception of sustainability: Do you think that this intervention should continue in your community? And extended in your country? Why?

###### If yes, do you see any conditions for it to be well implemented? How could the NutriVax strategy be adapted/scaled up to improve the access to vaccination in your community?
