## Supplementary File 4 for "Acceptability and feasibility of measles vaccination, small-quantity lipid-based nutrient supplementation, and their co-delivery among children aged 6–23 months in northern Nigeria: a qualitative study"

**NutriVax – Measles**

**Rapid summaries**

**Rapid summary - Parents/Guardian**

| **Access to Health Care** | | | |
| --- | --- | --- | --- |
| **Sub-Theme** | **Facilitators** | **Barriers** | **N**otes |
| **Experiences and logistical challenges** |  |  |  |
| **Decision-making** |  |  |  |
| **Quality of care and trust in services** |  |  |  |
| **Community dynamics** |  |  |  |
| **Vaccination** | | | |
| **Knowledge and perception** |  |  |  |
| **Vaccination status and decision-making** |  |  |  |
| **Community influences** |  |  |  |
| **Experiences of vaccination** |  |  |  |
| **Potential and additional motivators** |  |  |  |
| **SQ-LNS supplementation (only in the NutriVax arm)** | | | |
| **General Awareness and perception** |  |  |  |
| **Usage and adherence** |  |  |  |
| **Experiences and perception regarding distribution of SQ-LNS** |  |  |  |
| **Sustainability** | | | |
| **Perceptions regarding overall access to vaccination** |  |  |  |
| **Perceptions regarding the NutriVax strategy and sustainability** |  |  |  |

**Rapid summary – Community representatives**

| **Access to Health Care** | | | |
| --- | --- | --- | --- |
| **Sub-Theme** | **Facilitators** | **Barriers** | **N**otes |
| **General perceptions** |  |  |  |
| **Decision-making** |  |  |  |
| **Quality of care and trust in services** |  |  |  |
| **Community events** |  |  |  |
| **Vaccination** | | | |
| **Knowledge and perception** |  |  |  |
| **Parental reactions** |  |  |  |
| **Community events** |  |  |  |
| **SQ-LNS supplementation (only in the NutriVax arm)** | | | |
| **General Awareness and perception** |  |  |  |
| **Community reactions** |  |  |  |
| **External context** |  |  |  |
| **Relations with the** **NutriVax project team** | | | |
| **General interactions** |  |  |  |
| **Sustainability** | | | |
| **Perceptions regarding overall access to vaccination** |  |  |  |
| **Perceptions regarding the NutriVax strategy and sustainability** |  |  |  |

**Rapid summary – Community health workers**

| **Access to Health Care** | | | |
| --- | --- | --- | --- |
| **Sub-Theme** | **Facilitators** | **Barriers** | **N**otes |
| **General perceptions** |  |  |  |
| **Decision-making** |  |  |  |
| **Quality of care and trust in services** |  |  |  |
| **Community events** |  |  |  |
| **Vaccination** | | | |
| **Knowledge and perception** |  |  |  |
| **Parental reactions** |  |  |  |
| **Community events** |  |  |  |
| **SQ-LNS supplementation (only in the NutriVax arm)** | | | |
| **General Awareness and perception** |  |  |  |
| **Community reactions** |  |  |  |
| **Adherence and practices** |  |  |  |
| **Sharing practices** |  |  |  |
| **External context** |  |  |  |
| **Working conditions and relations with the** **NutriVax project team** | | | |
| **General perceptions** |  |  |  |
| **Collaborations and dynamics** |  |  |  |
| **Sustainability** | | | |
| **Perceptions regarding overall access to vaccination** |  |  |  |
| **Perceptions regarding the NutriVax strategy and sustainability** |  |  |  |

**Rapid summary – Healthcare workers**

| **Access to Health Care** | | | |
| --- | --- | --- | --- |
| **Sub-Theme** | **Facilitators** | **Barriers** | **N**otes |
| **General perceptions** |  |  |  |
| **Decision-making** |  |  |  |
| **Community events** |  |  |  |
| **Vaccination** | | | |
| **General perception** |  |  |  |
| **Parental reactions** |  |  |  |
| **Community events** |  |  |  |
| **SQ-LNS supplementation (only in the NutriVax arm)** | | | |
| **General Awareness and perception** |  |  |  |
| **Parental reactions** |  |  |  |
| **Adherence and practices** |  |  |  |
| **Sharing practices** |  |  |  |
| **External context** |  |  |  |
| **Working conditions and relations with the** **NutriVax project team** | | | |
| **General perceptions** |  |  |  |
| **Collaborations and dynamics** |  |  |  |
| **Sustainability** | | | |
| **Perceptions regarding overall access to vaccination** |  |  |  |
| **Perceptions regarding the NutriVax strategy and sustainability** |  |  |  |
