## Supplementary File 5 for "Acceptability and feasibility of measles vaccination, small-quantity lipid-based nutrient supplementation, and their co-delivery among children aged 6–23 months in northern Nigeria: a qualitative study"

**Table S1. Summary of access to care results analysed using the expanded Andersen’s Conceptual Framework and the Levesque Framework.**

| **Construct** | **Remaining facilitators** | **Remaining barriers** | **Impact of implementation adaptations** |
| --- | --- | --- | --- |
| **Need factors** (i.e. perceptions of the health status) | - Health education and mobilization activities by CNMs, HCWs, Community leaders, Ministry of Health (including targeted efforts for fathers) increase awareness on vaccination, and the importance of healthcare visits. - Disease outbreaks (measles, cholera, diphtheria) increase awareness and attendance. - Visible recovery of children brought to the hospital encourages parents to seek care. - Parents’ knowledge and recognition of measles risk. - Ongoing ANC sensitization. | - Preference for the intervention arm facilities over the control arms due to perceived benefits (SQ-LNS). - Some parents still delay care until session days. - Care-seeking only when illness is severe. |  |
| **Psychological factors – Social environment** (i.e. attitudes, knowledge, social norms, decision making) | - Support from village heads, community leaders, communal initiatives and interventions from other organizations. - SQ-LNS encourages adherence and wider service uptake. - Awareness creation and mobilization by community leaders using town criers. - Health awareness activities, Measles vaccine campaigns increase knowledge of available services. - Satisfactory and quality care, good waiting areas. - Joint healthcare decision-making by parents, with mothers often taking children to the facility and fathers providing financial and moral support. - Some households have fathers as primary decision-makers, while others allow mothers to decide independently. - No cultural or gender-based barriers to healthcare access. | - Vaccine rejection by some fathers and the prevailing social norm that mothers must obtain permission before seeking care. - Spousal disagreements influenced by social and religious norms requiring fathers’ approval in healthcare decision-making. - Beliefs linking certain diseases to supernatural causes. - Lack of awareness about health services in some areas. - Preference for specific Healthcare facility/HCW based on their attitude or service quality. - Belief in traditional medicine over orthodox healthcare among some community members. - Some parents to be attended to by healthcare workers from their own community - Some parents preferred to be attended to by healthcare workers from their own community. | - Male Sensitization improved gender norms around healthcare visits and vaccination. |
| **Enabling factors / Ability to reach** **& Ability to pay** (i.e. possibility of care access including availability - distance to health facilities, or financial resources) | - Nearby health facilities (PHCC, PHC and HP) - Decentralized RI/SQ-LNS distribution points. - Free medication when available through facilities or supporting NGOs. - 24-hour service availability (including ANC, SRH, emergency care) - Reliable HCW presence and minimal waiting times. - Adequate facility infrastructure: waiting areas, chairs, and sickbeds. | - Long distances to some facilities, poor road networks, especially during rainy season. - Lack of transportation options, high transport costs, and no ambulance services. - Flooding along access roads and within facilities, sometimes requiring relocation. - Curfews/protests disrupt access. - High out-of-pocket costs for healthcare (ANC, medication, transport). - Shortages of drugs and essential supplies in facilities. - Shortage of HCWs, leading to long waiting times. - Inadequate hospital space and water scarcity affecting operations. |  |
| **Psychological factors – health system** (i.e. perceptions regarding health facilities quality, relations with healthcare providers) | - Good and quality healthcare services (ANC and general care) - Good reception, attention, and care from HCWs (especially from older staff) - Availability of midwives. - Perceived better service organization | - Limited staff and space at some facilities. - Long waiting times during peak periods. | - Respectful HCW behavior encouraged parents’ attendance. - House-to-house visits with mobilizers |

**Table S2. Summary of vaccination results analyzed using the WHO Behavioural and Social Drivers of Vaccination framework**

| **Construct** | **Remaining facilitators** | **Remaining barriers** | **Impact of implementation adaptations** |
| --- | --- | --- | --- |
| **Thinking & feeling** (i.e. perceived disease risk, vaccine confidence) | - Health talks and sensitization before/after vaccination improve awareness. - Seeing the effectiveness of vaccines in preventing diseases like measles and whooping cough motivates uptake. - Seeing that measles cases are now rare makes parent belief in vaccine efficacy - Observed difference among vaccinated children and those not vaccinated - Fear of measles and its complications (e.g., blindness) motivates vaccination. - Concern over the high cost of treating vaccine-preventable diseases encourages prevention. - Good understanding of immunization schedules and processes, especially among literate parents. - Post-immunization health talk reduces fear and counters misinformation.   Awareness campaigns by CNMs, community leaders increase acceptance. | - Fear of adverse events following immunization (pain, fever, excessive crying - Doubts about effectiveness of vaccines - Misconceptions that vaccines can cause infertility. - Communication gaps where HCWs do not adequately explain vaccination importance. | - More systematic pre- and post-immunization health talks improved vaccination rates. - Collaboration with LGA officials to ensure a steady supply of paracetamol, as requested by some parents during the intermediate evaluation. - Increased awareness of post-immunization symptoms due to awareness campaigns. - Hiring local mobilizers for awareness campaigns |
| **Social processes** (i.e. social norms regarding vaccination, gender equity) | - Increased community mobilization and awareness activities, particularly involving fathers, men, and leaders, with sensitization by Community Nutrition Mobilizers (CNMs) boosting awareness and participation. - Peer-to-peer influence and sharing of positive experiences - Involvement of community leaders directly in awareness creation - Mobilizers recruited from within settlements - Male engagement encourage participation - Some fathers’ support for child vaccination. - Higher education levels influenced parents’ understanding of vaccine benefits. - Religious leaders’ involvement promotes acceptance.   Community leaders lead by example, supporting vaccination. | - Some did not support child vaccination (considering that fathers have a significant role in household healthcare decisions). - Some parents disregard CNMs during awareness sessions | - Increased Community sensitizations - Hiring local mobilizers improved trust. - Involving community & religious leaders - Deploy more CNMs on session days   Male resistance shifted to encouragement following targeted awareness campaigns directed at men. |
| **Motivation** (i.e. intention to get recommended vaccine) | - SQ-LNS distribution as incentives encourage adherence to the immunization schedule - Other incentives, such as mosquito nets, soap, and food items provided by NGOs, motivate vaccine uptake. - Knowledge of vaccine efficacy, particularly for measles improves uptake.   Spousal support and motivation encourage health-seeking behavior and participation in immunization. | - Forgetfulness of immunization schedule among parents.   Occasional barriers due to competing priorities like farming activities and market days. | Consistent community mobilization by HCWs and CHWs |
| **Practical issue** (i.e. availability, ease of access, service quality, respect from health workers) | - Consistent availability of vaccines, SQ-LNS, and vaccination materials. - Functional cold chain and good storage facilities - No shortages - Minimal waiting times in some facilities, sufficient manpower at vaccination points. - Availability of RI providers and free vaccination services. - Proximity of vaccination points improves access.   Good rapport and trust in CNMs and HCWs. | - Distance to facilities, poor road access, and seasonal flooding limiting access to vaccination services - Farming season reduces availability of parents. - Long waiting times in some facilities due to pooling of children before opening vaccine vials. - SQ-LNS ineligible parents refuse vaccination. - Preference for house-to-house vaccination for easier access.   Poor vaccine storage facilities in some facilities. | - Sharing of SQ-LNS at waiting area to make the children comfortable during waiting times - Improved waiting areas and ongoing health talks |

**Table S3. Summary of SQ-LNS supplementation results analyzed using Peh’s Adherence Framework.**

| **Construct** | **Remaining facilitators** | **Remaining barriers** | **Impact of implementation adaptations** |
| --- | --- | --- | --- |
| **Condition-related factors** (i.e. disease control and characteristics, patient specific) |  |  |  |
| **Medication-related factors** (i.e. medication regimen, medication effects, medication properties) | - SQ-LNS taste is acceptable and encourages adherence. - Children enjoy SQ-LNS, increasing compliance. - Small sachet design discouraged resale. - Benefits to child health/growth - Direct consumption of SQ-LNS from sachet or mixing with food   Perceived child strength, improved appetite, prevention of malnutrition, and mothers’ knowledge of SQ-LNS benefits motivated uptake. | - Complaints about the small size of SQ-LNS, as some parents perceived the quantity to be less than RUTF and therefore less effective. - Some caregivers shared SQ-LNS with ineligible children - Misuse of SQ-LNS incentives, including borrowing children and occasional sale. | Advocacy emphasizing that SQ-LNS should be given only to eligible children. |
| **Healthcare system factors** (i.e. healthcare providers characteristics (relationship, trust, training…), health facility access, drug supply, information support) | - Proximity of facility and decentralized RI/SQ-LNS points (PHCC). - Consistent vaccine/SQ-LNS availability and cold chain. - Regular staff training/supervision. - Good teamwork and coordination (facility, ALIMA, CNMs). - Mobilization of parents at community/facility levels. - Peer-to-peer information sharing. - Parents attend in groups for SQ-LNS collection. - Support from ALIMA through the NutriVax program (free medication). - Good attitudes, care, and courtesy from HCW - Good rapport and trust between HCWs, CNMs and parents. | - Late arrivals of some parents lead to missed services. - Long queues and waiting times. - Increased workload due to NutriVax turnout. - Parents dissatisfaction due to rejection of children without RI cards or late arrivals - Complaints about lack of additional support beyond vaccines and SQ-LNS. | - Provide multiple rations of SQ-LNS to parents from distance communities - Improved facility infrastructure (waiting area, shades, seats and mats). - Distribute one SQ-LNS sachet in waiting areas Service reorganization (early clinic opening) - Introduction of a new register improved workflow efficiency, reduced delays, and minimized administrative errors. - Deploy more CNMs on session days - Prioritize caretakers from far locations - Male engagement.   Positive attitude of HCWs and CNMs - staff are more respectful and organized. |
| **Patient-related factors** (i.e. cognitive and psychological factors, behavioral factors, priorities, non-modifiable characteristics, family/caregiver support) | - Spousal/household support for decisions and financing. - Peer influence among parents.   Parental literacy influences compliance. | - Some fathers unsupportive of mobilization. | Male engagement |
| **Socio-economic factors** (i.e. social/environmental context, norms, lifestyle factors, economic factors) | - Community leaders’ influence promotes participation. - Sharing of vaccine and SQ-LNS success stories as evidence. - Community mobilization and cultural acceptance of SQ-LNS noted. - Education on SQ-LNS at sessions/gatherings supports use   Improved work environment for beneficiaries and staffs enhances delivery. | - Transport costs still constrain some families | - Engagement of community and religious leaders - Giving monthly rations to distant parents to reduce travelling burden |

**Table S4. Summary of working condition** results analyzed using the inner setting the CFIR framework.

| **Construct** | **Remaining facilitators** | **Remaining barriers** | **Impact of adaptations suggested following the intermediate phase.** |
| --- | --- | --- | --- |
| **Workload** | - Increased patient flow and facility attendance due to the NutriVax intervention. - Staff ability to adapt to increased workload - Cooperation by parents during home visits and facility attendance. - Monthly incentives - Positive motivation of HCWs/CHWs - Team support | - Increased workload for health workers due to the NutriVax intervention and high patient turnout. - Shortage of staff at vaccination points. - Long waiting times at service points. - Limited staff for high workload - Extra time required on distribution days |  |
| **Relationship with colleagues** | - Good relationship, coordination, and collaboration between facility staff and ALIMA - Peaceful working relationships and good interactions within teams. - Regular supervision | - Occasional tension when resources are scarce | - New register improved workflow efficiency.  • Reduced delays and administrative errors. |
| **Work environment & resources** | - Adequate waiting areas, seating, shaded spaces, mats, and water supply. - Improved facility infrastructure following the NutriVax intervention. - Availability of vaccines, SQ-LNS, and provision of incentives. - Increased patient flow and increase in facility activities (e.g., immunization, ANC, OPD) due to NutriVax intervention and community mobilization. - Mobilization at both community and facility levels on health and hygiene - Improved work environment for staff. - Refresher training - Support from ALIMA (medication, nets) | - Long distance to facilities and poor road networks hindering access, especially for those outside the ward. - Staff mobility - Shortage of healthcare workers. - Rejection of children without RI cards or those arriving late. - Late coming for non-resident staff |  |
